# HIV infection is linked with reduced error-related default mode network suppression and poorer medication management abilities

**DOI:** 10.1101/2021.04.10.21255223

**Authors:** Jessica S. Flannery, Michael C. Riedel, Taylor Salo, Ranjita Poudel, Angela R. Laird, Raul Gonzalez, Matthew T. Sutherland

## Abstract

**Objective:** Brain activity linked with error processing has rarely been examined among persons living with HIV (PLWH) despite importance for monitoring and modifying behaviors that could lead to adverse health outcomes (e.g., medication non-adherence, drug use, risky sexual practices). Given that cannabis (CB) use is prevalent among PLWH and impacts error processing, we assessed the influence of HIV serostatus and chronic CB use on error-related brain activity while also considering associated implications for everyday functioning and clinically-relevant disease management behaviors.

**Methods:** A sample of 109 participants, stratified into four groups by HIV and CB (HIV+/CB+, *n*=32; HIV+/CB-, *n*=27; HIV-/CB+, *n*=28; HIV-/CB-, *n*=22), underwent fMRI scanning while completing a modified Go/NoGo paradigm called the Error Awareness Task (EAT). Participants also completed a battery of well-validated instruments including a subjective report of everyday cognitive failures and an objective measure of medication management abilities.

**Results:** Across all participants, we observed expected error-related anterior insula (aI) activation which correlated with better task performance (i.e., less errors) and, among HIV-participants, fewer self-reported cognitive failures. Regarding awareness, greater insula *activation* as well as greater posterior cingulate cortex (PCC) *deactivation* were notably linked with aware (vs. unaware) errors. Regarding group effects, unlike HIV-participants, PLWH displayed a lack of error-related deactivation in two default mode network (DMN) regions (i.e., PCC, medial prefrontal cortex [mPFC]). No CB main or interaction effects were detected. Across all participants, reduced error-related PCC deactivation correlated with reduced medication management abilities and PCC deactivation mediated the effect of HIV on such abilities. More lifetime CB use was linked with reduced error-related mPFC deactivation among HIV-participants and poorer medication management across CB users.

**Conclusions:** These results demonstrate that insufficient error-related DMN suppression linked with HIV infection, as well as chronic CB use among HIV-participants, has real-world consequences for medication management behaviors. We speculate that insufficient DMN suppression may reflect an inability to disengage task irrelevant mental operations, ultimately hindering error monitoring and behavior modification.

## INTRODUCTION

HIV crosses the blood brain barrier, replicates, and infects cells in the central nervous system (CNS), leading to neuroinflammation and neural degeneration [1-3]. Neuroimaging has identified structural and functional brain alterations linked with HIV infection, particularly in frontostriatal circuitry [4-9], that are thought to contribute to a spectrum of progressive neurocognitive declines characterizing the disease [10-15]. Even with widespread availability of antiretroviral therapy, approximately 30–50% of persons living with HIV (PLWH) are impacted by neurocognitive alterations [16, 17]. Accumulating evidence indicates that PLWH not only exhibit altered neurocognition but also manifest metacognitive difficulties whereby they under report their cognitive failures when considered in light of objective behavioral measures [18-21]. Reduced awareness of everyday mistakes and errors may impact daily functioning, potentially resulting in compromised medication adherence and, in turn, disease management.

Highly active antiretroviral therapies (HAART) suppress viral load, improve cognitive function [22], and prolong life for PLWH [23, 24]. Yet, strict medication adherence is critical for HAART success [24-27] with even a few days of missed doses leading to viral load increase [28, 29] and drug resistance [30, 31]. HAART dosing regimens are also complicated, demanding, and challenging to manage; for example, use of a protease inhibitor requires dosing every 8-or 12 hours and at least two additional antivirals on potentially different schedules [31]. Cognitive control, attentional, and psychomotor impairments linked with the infection likely make managing complex medication regimens even more challenging [31, 32]. Consequently, medication adherence is a major barrier to sustained health for PLWH [22] and adherence rates are estimated to only reach 40-67% [33]. Monitoring and recognizing cognitive failures is likely relevant to medication management as it allows one to rectify errors and adapt behavior thereby avoiding future negative outcomes.

Given recent changes in societal views, state laws, and clinical practice, recreational and medical cannabis (CB) use remains prevalent among PLWH [34-36] with 77% reporting lifetime use [37]. Despite anti-inflammatory and other benefits (e.g., analgesic, gastrointestinal) potentially linked with CB use by PLWH [36, 38], evidence suggests chronic use may lead to neurocognitive alterations [39-41]. Such alterations appear dependent on CB use histories (e.g., duration, amount [40, 42-44]) and possibly differ across PLWH and HIV-users [40, 42]. However, given inconsistencies in the literature, the neuroprotective versus potential adverse synergistic impact of CB use and HIV infection remains unclear [41] as does potential consequences for complex real-world behaviors such as medication management [45, 46].

There is growing appreciation that neurocognitive behavioral measures alone may not provide a complete perspective on the potential neurobiological impacts of HIV and CB [41, 47]. Indeed, atypical brain activity among PLWH, measured via functional magnetic resonance imaging (fMRI) may precede observable declines in cognition and diagnoses of HIV-associated neurocognitive disorder (HAND) [47]. Although medication management abilities have been linked with neurocognitive dysfunction among PLWH [31], altered brain activity underlying and perhaps predating such dysfunction remains to be characterized. Here, we considered how brain activity linked with cognitive control, error processing, and error awareness may be altered as both a function of HIV infection and CB use in the service of enhancing insight into the neurobiological mechanisms potentially contributing to medication adherence difficulties.

Accumulating neuroimaging evidence implicates both HIV and CB when considering functional alterations in brain regions contributing to error processing and awareness. For example, elevated task-related insula activity among PLWH [48] and reduced error awareness-related insula and dorsal medial prefrontal cortex (dmPFC) activity among CB users [49] has been documented. Critically, HIV x CB interactive effects on left anterior insula (aI) activity have been reported in the context of a cognitive interference fMRI task, such that CB use was associated with increased aI activity among PLWH, yet decreased activity among HIV-participants [50]. The aI and dmPFC are central nodes of the salience network (SN), are critically involved in monitoring errors and, more generally, salient stimuli, as well as the deployment of attentional resources [51-54]. SN regions are also implicated in explicit error awareness [51, 52, 55, 56], are relevant for optimal task performance [49, 57], and are linked with subjective awareness of cognitive deficits (i.e., insight) across neuropsychiatric conditions [55, 58, 59]. Another large-scale brain network potentially relevant for optimal cognition is the default mode network (DMN). The posterior cingulate cortex (PCC) and medial prefrontal cortex (mPFC) are central DMN nodes [60-62], regions thought to be more active during task irrelevant mental operations and to deactivate during “task-on” performance [60, 63]. As a robust body of work has shown that trial-by-trial DMN suppression is related to optimal task performance [64, 65], detection of salient stimuli [66], and increasing cognitive demands [63, 67], DMN suppression is also likely relevant in the context of error processing. While insufficient DMN suppression has been implicated in various neuropsychiatric disorders [53, 68-76], it remains to be considered among PLWH.

As such, we probed brain activity linked with cognitive control (e.g., inhibition), cognitive failures (i.e., error commission), and explicit error awareness utilizing a Go/NoGo motor inhibition paradigm called the Error Awareness Task (EAT) among a participant sample stratified by HIV serostatus and CB use history. We examined the potential interactive impacts of HIV and CB on brain responsivity to task errors, of which participants were either aware or unaware. Importantly, we also considered clinically-relevant implications by delineating relationships between brain activity and behavioral measures of cognitive control, error awareness, and medication management ability, as well as self-reported everyday cognitive failures. We addressed three main empirical questions involving task-effects, group-effects, and real-world implications. Regarding task effects, we expected to replicate *error-related* activity in SN regions and previously reported *awareness-related* brain activations (SN regions) and deactivations (DMN regions). Regarding group effects, we anticipated observing HIV x CB interactive effects on EAT-related SN and DMN regional brain activity. Finally, regarding real-world implications, we expected that error-and/or awareness-related brain activity showing group differences would be linked with subjectively reported cognitive failures and objectively measured medication management abilities.

## METHODS

### 2.1. Participants

A sample of 109 participants was stratified into four groups based on HIV serostatus and CB use history (HIV+/CB+, *n*=32; HIV+/CB-, *n*=27; HIV-/CB+, *n*=28; HIV-/CB-, *n*=22). Participants were recruited from community-based organizations providing health care services throughout Miami-Dade County. All participants were 18-60 years old to minimize the presence of other chronic conditions (e.g., hypertension, diabetes), as well as the potential interactive effect of HIV and aging on neurocognition [77-81]. Additional exclusionary criteria included: current Hepatitis C infection, English non-fluency or illiteracy, less than an eighth-grade education level, severe learning disability, serious neurological disorder, severe head trauma with loss of consciousness >30 min, severe mental illness with psychotic or paranoid symptoms, or MRI contraindications. All PLWH in the study were taking antiretroviral medications, were diagnosed with HIV 9.8±9.2 (mean±SD) years prior to assessment, and had no history of opportunistic infections affecting the CNS. All CB using participants reported a history of regular use (at least once per week for three straight weeks) and used at least 20 times in the past year. CB-participants met the following criteria: no CB use in the past 12 months, and a negative urine THC screen. Past use of and dependence on other substances, including alcohol, nicotine, cocaine, amphetamines, benzodiazepines, or opioids was permitted across each group to provide a more representative and generalizable sample. However, participants were excluded if meeting criteria for current substance dependence (except nicotine and CB) as assessed via the DSM-5 Structured Clinical Interview [82].

### 2.2. Procedures

Study procedures were reviewed and approved by FIU’s Institutional Review Board. Following informed consent, we collected blood, behavioral, self-report, and MRI data across two study visits on different days. CB+ participants were instructed to refrain from use for 24 hours before study visits to minimize acute pharmacological and/or withdrawal-related effects. Upon arrival at both visits, participants completed substance use screening including urine toxicology (Drug Check Cup, NXStep) and breathalyzer testing (AlcoMate Premium Breathalyzer). During the first visit, blood specimens were collected and participants completed a battery of behavioral tests and self-report questionnaires. Among PLWH, blood samples were used to quantify HIV disease severity (HIV-1 viral load) and immune function (lymphocyte T-cell subset counts). Among CB+ participants, samples were used to quantify cannabinoid levels (plasma 9-carboxy-THC and THC to creatine ratios). The second visit occurred within 1 month after the first and participants completed a 1-hour MRI scan after task training and completion of additional self-reports. Participants were compensated at the end of each visit.

### 2.3. Behavioral and self-report measures

Participants completed a battery of well-validated behavioral and self-report instruments. Herein, we focus on measures pertaining to self-reported cognitive failures, medication management abilities, and detailed CB use history. To quantify self-awareness of everyday cognitive failures, we considered participants’ total scores on the Cognitive Failures Questionnaire (CFQ) [83]. The CFQ is a 25-item questionnaire assessing the occurrence of absent-mindedness or slips of perception, memory, and motor functioning. Among healthy participants, CFQ total scores are linked with inattentiveness, forgetfulness, and/or accidents in both the laboratory and real-world resulting from distractibility, poor selective attention, or other mental errors [84]. To quantify medication management abilities, participants completed the Revised Medication Management Test (MMT-R), a 10-minute behavioral test validated for use among PLWH [85]. The MMT-R assesses an individual’s ability to accurately follow a fictitious prescription regimen and answer questions about the mock medications [86, 87]. The MMT-R involves a pill dispensing component designed to emulate the complex, multi-medicine regimens currently used for HIV treatment and a medication inference component [87]. During the pill dispensing component, participants transferred mock pills from bottles to a 1-week pill organizer. To quantify CB use history, self-reported information on frequency, amount, and duration of CB (and other drug) use was collected via selected items from the National Survey on Drug Use and Health [88].

### 2.4. Error Awareness Task (EAT)

During MRI scanning, participants completed a modified Go/NoGo motor inhibition paradigm called the Error Awareness Task (EAT) [49, 89-91]. In the EAT, participants commit errors (i.e., incorrectly press a button following a NoGo cue) of which they can be either aware or unaware. Participants indicate error awareness by pressing an error signaling button. The EAT allows for assessment of distinct brain activity linked with cognitive control (e.g., inhibition), cognitive failures (i.e., error commission), and explicit error awareness.

During the task, participants viewed a series of color words (e.g., “RED”) presented one at a time which were written in a font/ink color that was either congruent with the word’s meaning (e.g., the word “RED” written in red font) or incongruent (e.g., “RED” in blue font). Words were initially displayed for 900ms followed by a 600ms inter-stimulus interval. Participants were instructed to press button-1 on a two-button response box (Current Designs, Philadelphia, PA) as quickly as possible during the stimulus display window each time a new word appeared (Go trials). Participants were instructed to withhold this button press under two conditions (NoGo trials). The first NoGo condition was when the same congruent word-ink stimulus was repeated on two consecutive trials (NoGo: repeat). The second was when the word’s meaning and ink color were incongruent (NoGo: Stroop). Go trials occurred more frequently than NoGo trials (5:1 ratio) to render a Go button press a prepotent response. An equal number of NoGo: repeat and NoGo: Stroop trials were pseudo-randomly presented across the task, such that there were at least 3 and at most 7 Go trials between each NoGo cue. The two NoGo conditions were included to increase task difficulty and ensure a sufficient number of errors [89]. Specifically, given the overlearned human tendency to read words relative to identifying the font color, participants were expected to more successfully monitor for the NoGo: repeat condition leading to fewer errors relative to the NoGo: Stroop condition. To indicate explicit awareness following commission errors, participants were trained to press an error signaling button (button-2) on the Go trial immediately following the erroneous response (hereafter referred to as the awareness trial). Successful inhibition in the EAT necessitates decision-making and multiple cognitive control processes. Specifically, identification of NoGo: repeat trials requires working memory maintenance of the preceding word whereas identification of NoGo: Stroop trials involves cognitive interference from the prepotent word reading response [89, 92]. Consequently, successful inhibition necessitates the execution of multiple, confounded, cognitive control processes including: sustained attention for monitoring stimuli, successful NoGo stimulus detection, and ultimately motor inhibition of the prepotent Go response. Thus, we considered successful motor inhibition as the culmination of a series of successful goal-directed cognitive control processes (as opposed to isolation of the response inhibition construct) and error commission as a general cognitive failure.

Participants completed EAT training and then performed a 5.5min practice run in a mock scanner. During MRI scanning, participants completed six, 5.5min task runs involving a total of 1,296 trials (1,080 Go trials, 216 NoGo trials [NoGo: repeat, *n*=108; NoGo: Stroop, *n*=108]) with short rest periods (∼30sec) between runs 1-2, 3-4, and 5-6 and longer breaks between runs 2-3 (6min for a structural MRI scan) and runs 4-5 (8min for a resting-state fMRI scan). To achieve sufficient numbers of successful and unsuccessful NoGo trials, task difficulty was individually and dynamically adapted to maintain participants’ average NoGo error rate between 45-50%. Specifically, after the first 40 trials, when a participant’s average NoGo error rate fell below 45%, the length of the response window (initially 900ms) was reduced by 250ms (minimum allowable: 500ms). Alternatively, when the average NoGo error rate rose above 50%, the response window increased by 250ms (maximum allowable: 1000ms).

### 2.5. EAT behavioral measures: Statistical analyses

Behavioral variables of interest included the: **a)** number of Go trials with a correct (Go-correct) or missed response (Go-error [omission]), **b)** number of successfully (NoGo-correct) and unsuccessfully inhibited NoGo trials (NoGo-error), and **c)** number of NoGo-errors for which participants indicated error awareness (NoGo-error: Aware), failed to indicate awareness (NoGo-error: Unaware), or did not respond on the awareness trial (NoGo-error: No response). Regarding response times (RTs), variables of interest included: average RT on Go trials immediately preceding (pre-error) and following (1-post-error) aware and unaware NoGo-errors. Given that the 1-post-error trial (i.e., the awareness trial) was confounded with the additional error signaling process, we also considered RTs on the subsequent Go trial (2-post-error). As the EAT’s difficulty manipulation was intended to maintain error rates at ∼45-50%, we did not anticipate between-group differences on this variable. However, we aimed to replicate prior reports of higher error rates for NoGo: Stroop versus NoGo: repeat trials (e.g., [49, 90]). As such, we performed a 3-way, 2(HIV: + vs. -) x 2(CB: + vs. -) x 2(NOGO-TYPE: Stroop vs. repeat) mixed-effects ANOVA focusing on the NOGO-TYPE main effect. To assess potential between-group differences in error awareness, we performed a similar 3-way ANOVA (2[HIV] x 2[CB] x 2[AWARENESS: aware vs. unaware]). To characterize RT patterns following aware and unaware errors (i.e., post-error speeding or slowing), we performed a 4-way, 2(HIV) x 2(CB) x 2(AWARENESS) x 3(TRIAL: pre-vs. 1-post-vs. 2-post-error) mixed-effects ANOVA. We focused on the AWARENESS x TRIAL interaction as we aimed to replicate RT reductions following aware (post-error speeding), but not unaware errors [89]. Behavioral data were analyzed with SPSS (v.26) and Python (2.7.10).

#### 2.6.1. MRI data acquisition and analysis

MRI data were collected on a GE Healthcare Signa MR750, 3-Tesla scanner with 32-channel head coil. For the six functional EAT runs, 42 slices (3.4mm thick) were obtained in the axial plane using a T2*-weighted, single-shot, gradient-echo, echo-planar imaging (EPI) sequence sensitive to blood oxygenation level–dependent (BOLD) effects (169 volumes/run, repetition time [TR]=2000ms, echo time [TE]=30ms, flip angle [FA]=75°, field of view=220mm, 64×64 matrix, voxel size = 3.44 × 3.44 × 3.40 mm^3^). These same EPI parameters were used to collect an 8min resting-state scan with eyes closed (245 volumes, data not reported herein). T1-weighted structural images were obtained using a magnetization-prepared rapid gradient-echo (MPRAGE) sequence (TR=2500ms; TE=3.7ms; FA=12°; voxel size=1mm^3^).

MRI data preprocessing was performed with FMRIPREP v1.1.1 [93], a Nipype-based tool [94] often employing Nilearn [95]. T1-weighted structural volumes were corrected for intensity non-uniformity (N4BiasFieldCorrection v2.1.0) [96] and skull-stripped (antsBrainExtraction.sh v2.1.0). Nonlinear registration (ANTs v2.1.0) was performed to spatially normalize T1-weighted volumes to the ICBM-152 asymmetrical template v2009c [97]. Functional data were motion corrected using MCFLIRT (FSL v5.0.9) [98] and slice-time corrected to the middle of each TR using 3dTshift (AFNI v16.2.07) [99]. Distortion correction was performed by co-registering functional images to corresponding, intensity inverted [100, 101] anatomical volumes constrained by an average field map template [102]. Functional images were then co-registered to corresponding T1-weighted volumes using boundary-based registration [103] with 9 degrees of freedom via bbregister (FreeSurfer v6.0.1). The motion correction transformations, distortion correction warp, functional-to-anatomical transformation, and anatomical-to-template warp were all concatenated and applied in a single step using Lanczos interpolation (antsApplyTransforms ANTs v2.1.0).

Subject-and group-level fMRI analyses were performed in AFNI (http://afni.nimh.nih.gov/afni/). Following preprocessing, the six EAT runs were smoothed to 8mm FWHM (3dBlurToFWHM) and time series were scaled to the voxel-wise mean (3dcalc) thereby allowing regression (*β*) coefficients, calculated per regressor and participant, to be interpreted as an approximation of percent BOLD signal change (% BOLD Δ) [104] from the implicit baseline. The first five functional volumes of each run and those with framewise displacement greater than 0.35mm were censored (1.2±3.9% of TRs). Groups did not differ in the number of censored TRs (HIV: *F*[*1, 105*]=0.6, *p*=0.4; CB: *F*[1, *105*]=1.9, *p*=0.2; HIVxCB: *F*[1, *105*]=0.7, *p*=0.4). Functional data were then entered into two separate subject-level general linear models (GLMs) including task-related and nuisance regressors (i.e., six motion-correction parameters and fourth-order polynomials capturing residual head motion and baseline trends in the BOLD signal, respectively). To characterize brain activity linked with **cognitive control/failures**, the first GLM included three task-related regressors (NoGo-correct [C], NoGo-error [E], and Go-error [O, omission]) as impulse functions time-locked to stimulus onset and convolved with a hemodynamic response (gamma) function. To characterize brain activity linked with **error awareness**, the second GLM included the same nuisance regressors and now four task-related regressors again including NoGo-correct [C] and Go-error [O] regressors, but here, NoGo-errors [E] were parsed into two types, 1) NoGo-error: Aware [A], and 2) NoGo-error: Unaware [U].

#### 2.6.2. EAT-related brain activity: Task effects

Task-and group-effect statistical tests were conducted within a sample-specific whole-brain mask that included those voxels in which ≥80% of participants’ functional runs had a non-zero value. This overlap mask’s perimeter was eroded by one voxel yielding a final mask with 41,726 voxels. To characterize **cognitive control/failure**-related activity, NoGo-correct minus NoGo-error [C-E] contrast values were assessed across a sample of 103 participants demonstrating sufficient task engagement (i.e., responded to >50% of Go trials) with a whole-brain, one-sample *t-*test (two-tailed, 3dTtest++). For visualization, the resulting maps were thresholded at *p*_*voxel-wise*_=1.0e^-10^, cluster extent: 20 voxels (arbitrarily chosen). To assess **error awareness**-related activity, a whole-brain, one-sample *t-*test on NoGo-error: Aware minus NoGo-error: Unaware [A-U] contrast values was conducted across a sample of 86 participants demonstrating sufficient trial numbers (i.e., committed ≥2 aware and ≥2 unaware NoGo-errors) (*p*_*FWE-corrected*_ <0.05; *p*_*voxel-wise*_<0.0001, cluster extent: 7 voxels, 3dClustSim with spatial autocorrelation correction [105]).

For graphical examination and follow-up analyses, we extracted the average *β* coefficients associated with specific task events ([C], [E], [A], [U]) and contrasts ([C-E], [A-U]) by averaging across all non-zero voxels within clusters/regions of interest (ROIs) identified via the **cognitive control/failure** and **error awareness** statistical maps. To assess brain-behavior relationships, we considered the total number of NoGo-errors (ERROR COUNT) as an objective, laboratory-based measure of cognitive failures and the total scores on the CFQ (CFQ) as a subjective measure of real-world cognitive failures. We also considered the percent of unaware NoGo-errors (% UNAWARE) as an objective measure of error awareness. First, we performed bivariate Pearson’s correlations between the average **cognitive control/failure** error-specific *β* coefficients [E] and ERROR COUNT. These analyses were Bonferronni-corrected for the *n*=4 ROIs considered. Regarding **error awareness**-related activity, we performed correlations between [A-U] contrast values and % UNAWARE (Bonferronni-corrected, *n*=4). As the % UNAWARE distribution was right-skewed, this variable was log_10_ transformed. Additionally, we conducted exploratory analyses examining whether HIV serostatus moderated the relationship between self-reported cognitive failures (CFQ) and behavioral performance (ERROR COUNT, % UNAWARE) or task-related brain activity (error [E] *β* coefficients, [A-U] contrast values) by conducting separate HIV x CFQ ANCOVAs. As these analyses were exploratory, we did not apply multiple comparison corrections.

#### 2.6.3. EAT-related brain activity: Group effects

Group differences in **cognitive-control/failure** [C-E] and **error awareness** [A-U] activity were assessed via whole-brain, 2(HIV) x 2(CB) ANOVAs (3dMVM). HIV and CB main and interaction effects were of interest (*p*_*FWE-corrected*_<0.05; *p*_*voxel-wise*_<0.001, cluster extent: 35 voxels). These group-effects analyses were also performed using sex, age, and IQ as covariates. No significant covariate influences were detected; thus, results from the initial analyses are reported. The *β* coefficients and contrast values from identified ROIs were extracted for graphical examination and follow-up analyses. We assessed whether HIV serostatus moderated the relationship between behavioral performance (ERROR COUNT) and task-related brain activity (error [E] *β* coefficients) by conducting HIV x ERROR COUNT ANCOVAs (Bonferroni-corrected for *n*=2 ROIs considered).

To more fully characterize group differences in brain activity across various trials, we computed *β* coefficients for the three Go trials both preceding (3-pre, 2-pre, 1-pre) and following a NoGo-error (1-post [awareness trial], 2-post, 3-post). Specifically, we performed six separate GLMs similar to that described above when considering cognitive control/failures, except in these GLMs, the NoGo-error [E] regressor was replaced with one modeling a specific error-preceding or error-succeeding Go trial. *β* coefficients were extracted from ROIs and plotted with respect to NoGo-error [E] and NoGo-correct [C] activity for qualitative inspection and exploratory analyses. We performed a 2(HIV) x 6(TRIAL: 3-pre vs. 2-pre vs. 1-pre vs. 1-post vs. 2-post vs. 3-post) mixed-effects ANOVA on each ROI’s *β* coefficients with a focus on the HIV x TRIAL interaction (Bonferroni-corrected for *n*=2 ROIs).

To probe clinically-relevant implications of HIV-associated alterations, we examined relationships between error-related brain activity, behavioral performance on the MMT-R (MMT SCORE), and self-reported CFQ total scores (CFQ). Specifically, we conducted HIV x MMT SCORE and HIV x CFQ ANCOVAs when considering NoGo-error activity [E] extracted from those ROIs demonstrating group effects above. Follow-up conditional effects within groups were assessed using the PROCESS v.5 SPSS plugin [106]. Finally, we tested a mediation model (corresponding to Model 4 in Hayes, 2017 [106]) in which error-related brain activity (M) mediated the effect of HIV serostatus (X) on MMT SCORE (Y). While the cross-sectional and observational nature of our research design limits conclusions about causality, we aimed to provide a descriptive (as opposed to predictive) account of hypothesized co-relations between these variables [106, 107]. Finally, to evaluate the impact of self-reported lifetime CB use (LIFETIME AMOUNT) among HIV+ and HIV-CB using participants (*n*=55), we assessed HIV x LIFETIME AMOUNT interactions (controlling for past month nicotine use) on task-related brain activity (error [E] *β* coefficients), MMT SCORE, and CFQ.

## RESULTS

### 3.1. Group characteristics

Groups did not differ in age, education, or IQ when considering main and interaction effects in HIV x CB ANOVAs (**Table 1**; *p*’s>0.1) nor in terms of race, ethnicity (**Table 1**; *p*’s>0.1), or history of major depressive episodes (**Supplemental Table S1**; *p*’s>0.6) when considering Chi-square tests comparing HIV+ vs. HIV-and CB+ vs. CB-groups. However, the HIV+ groups included a higher percentage of males (79.6% male) than the HIV-groups (54.0% male; *χ*^2^[*1,109*]=8.2, *p*=0.004), consistent with national estimates regarding the sex distribution (81% male) of new HIV diagnoses [105]. This difference was driven by the female/male composition among the CB+ groups (HIV+/CB+ vs. HIV-/CB+: *χ*^2^[*1,109*]=8.9, *p*=0.003), but not the CB-groups (HIV+/CB-vs. HIV-/CB-: *χ*^2^[*1,109*]=1.4, *p*=0.2).

**Table 1.**
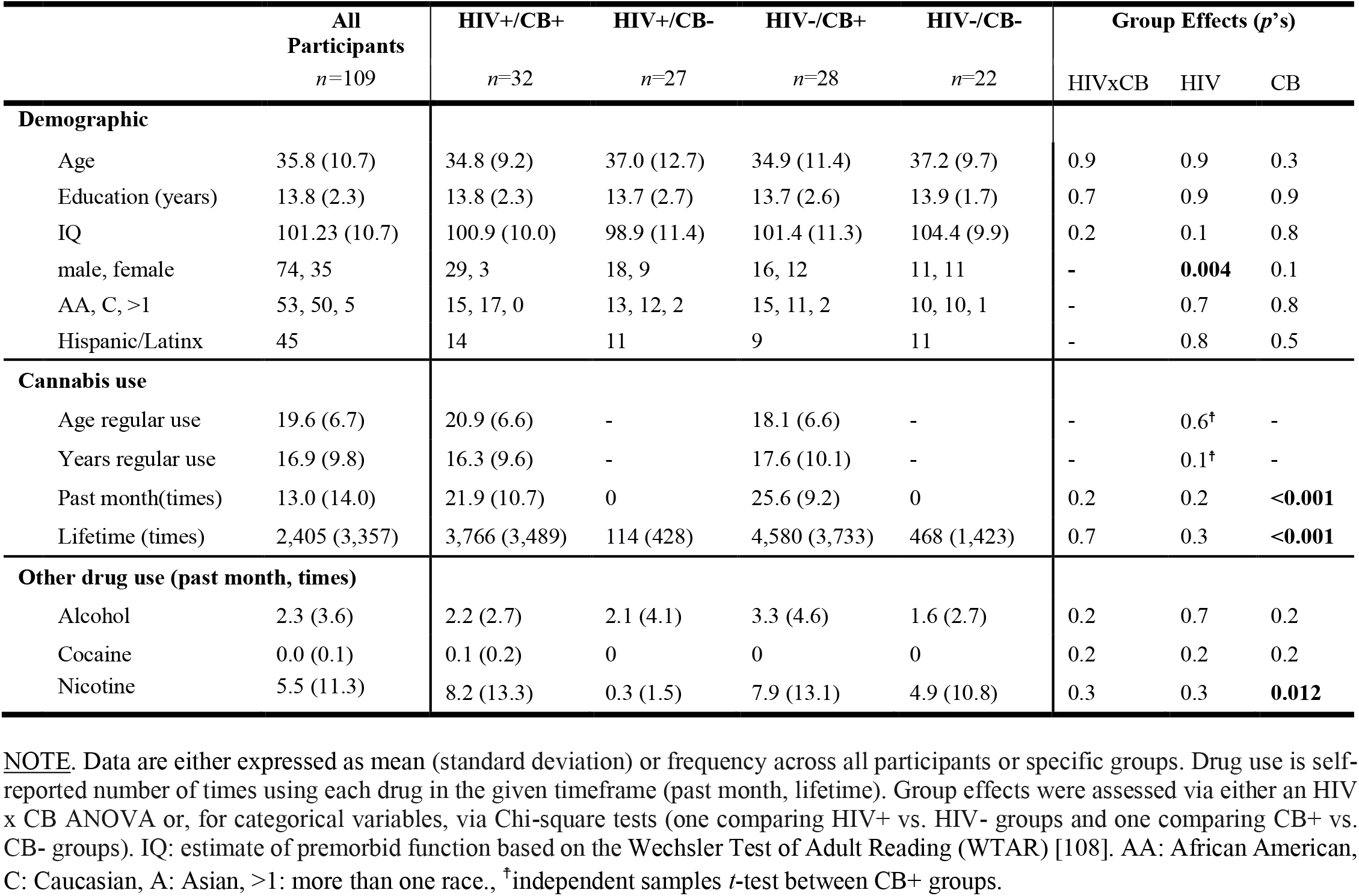
Participant demographic, cannabis use, and other drug use characteristics by group.

Groups composed of PLWH (HIV+/CB+ vs. HIV+/CB-) were largely matched on relevant measures of HIV-status (i.e., time since diagnosis, plasma viral load, % with detectable viral load, % diagnosed with AIDS), and immune function (i.e., lymphocyte T-cell subset counts, **Supplemental Table S2**; *p*’s>0.2). Similarly, the CB using groups (HIV+/CB+ vs. HIV-/CB+) were matched on self-reported measures of CB exposure (i.e., use duration, past month use, lifetime use, **Table 1**; *p*’s>0.1) and plasma measures of THC and metabolites (**Supplemental Table S3**; *p*’s>0.1). As expected, the CB+ groups had a higher percentage of participants meeting criteria for past (χ^2^[*1,109*]=12.1, *p*<0.001) and current (*χ*^2^[*1,109*]=10.0, *p*<0.002) CB dependence (**Supplemental Table S4**) and self-reported more past month (*F*[*1,109*]=52.9, *p*<0.001) and lifetime CB use (*F*[*1,109*]=269.5, *p*<0.001; **Table 1**).

Groups were largely matched on other drug use characteristics including past dependence (e.g., alcohol, cocaine; **Supplemental Table S4**) as well as past month and lifetime use (e.g., alcohol, cocaine, methamphetamine, prescription stimulants, heroin, opiates, benzodiazepines, barbiturates, inhalants; **Supplemental Table S5**). The only exceptions were nicotine and ecstasy, where the CB+ groups reported more past month nicotine use (*F*[*1,109*]=6.6, *p*=0.012) and more lifetime ecstasy use (*F*[*1,109*]=7.7, *p*=0.007; **Supplemental Table S5**).

### 3.2. EAT behavioral measures

Six participants (*n*=2 HIV+/CB+, *n*=2 HIV+/CB-, *n*=2 HIV-/CB+, *n*=0 HIV-/CB-) with more than 50% Go-errors (i.e., omissions) were excluded from subsequent behavioral and neuroimaging analyses. The remaining 103 participants, failed to respond on 3.1±0.6% (mean±SEM) of Go trials and failed to withhold a button press response on 46.1±1.5% of NoGo trials (**Table 2**). Given the dynamic task-difficulty manipulation, we did not observe any significant group effects when considering NoGo-errors in a 2(HIV) x 2(CB) x 2(NOGO-TYPE: Stroop vs. repeat) ANOVA (*p*’s>0.4). However, a NOGO-TYPE main effect was detected (*F*[*1,99*]=85.5, *p*<0.001) such that more errors were committed on Stroop (59.1±1.0% of NoGo-errors) versus repeat trials (40.9±1.0%, **Table 2, Fig. 1A**), consistent with previous reports (e.g. [49, 90]) and the interpretation that the NoGo: Stroop rule was more difficult to monitor [89].

**Table 2.** EAT behavioral performance measures.

|  | Trial Type | Mean ± SEM | Range |
| --- | --- | --- | --- |
| a) | % Go-correct | 96.9 ± 5.8 | 65.3 - 100 |
|  | % Go-error (omission) | 3.1 ± 0.6 | 0 - 34.7 |
| b) | % NoGo-correct | 53.9 ± 1.5 | 13.4 - 91.7 |
|  | % NoGo-error (commission) | 46.1 ± 1.5 | 8.3 - 86.6 |
| c) | % NoGo-error: Aware | 73.4 ± 2.4 | 0 - 100 |
|  | % NoGo-error: Unaware | 21.2 ± 2.3 | 0 - 100 |
|  | % NoGo-error: No response | 5.4 ± 0.7 | 0 - 33.5 |
| d) | % NoGo-error: Repeat | 40.9 ± 1.0 | 19.8 - 74.6 |
|  | % NoGo-error: Stroop | 59.1 ± 1.0 | 25.4 - 80.2 |

**NOTE.** Values in section **a)** are expressed as the percentage of trials out of all Go trials. Values in section **b)** are expressed as the percentage of trials out of all NoGo trials. Values in sections **c)** and **d)** are expressed as the percentage of trials out of all NoGo-errors. Values in each section (**a-d**) sum to 100%. SEM: standard error of the mean. See **Supplemental Table S6** for all EAT trial type counts and percentages.

**Figure 1.**
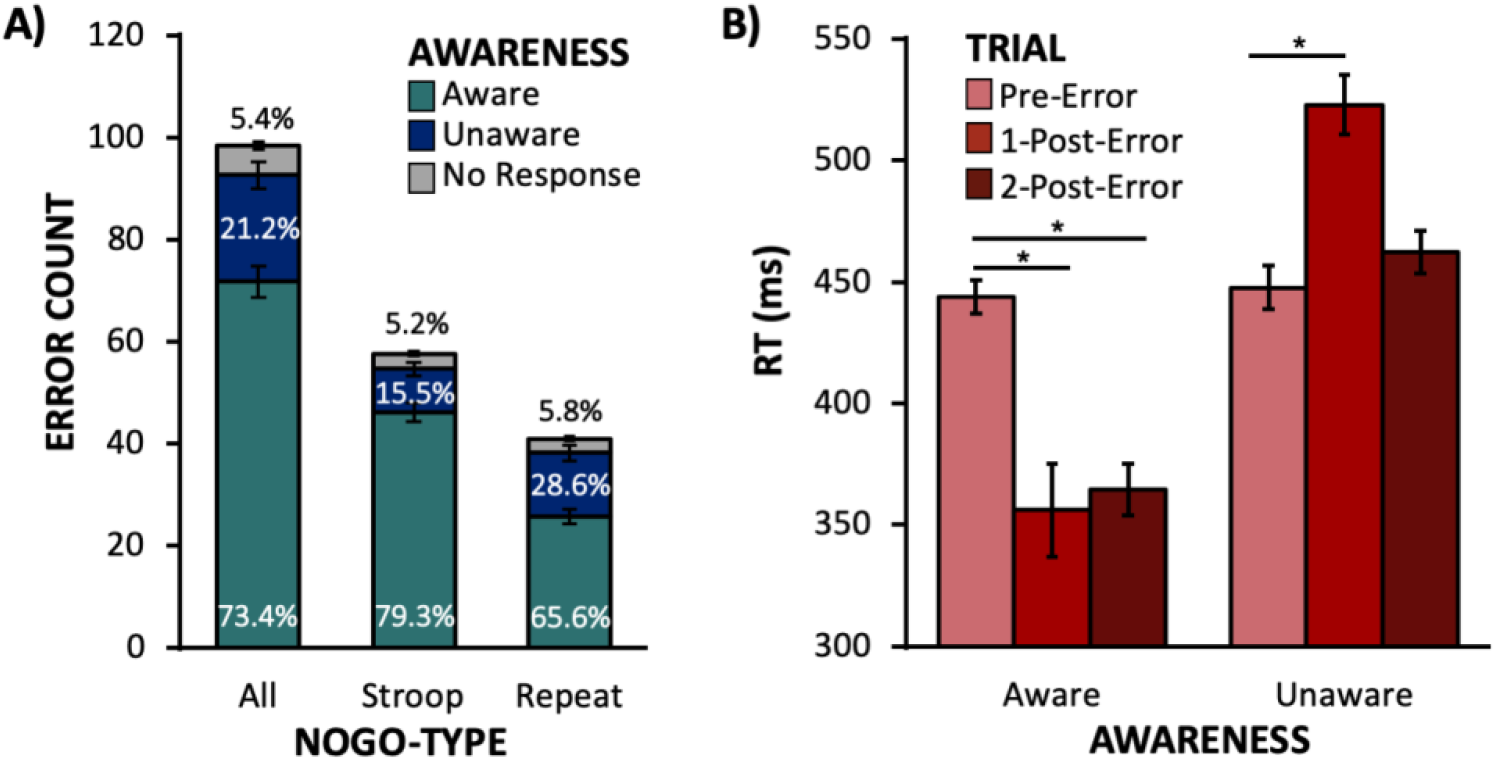
EAT behavioral performance measures. <u>**A)**</u> Participants indicated awareness of most errors (aqua: 73.4±2.4%) and were more likely to be aware of *Stroop errors* (79.3±2.3%) relative to *repeat errors* (65.6±2.8%; *t*[*102*]=7.7, *p*<0.001). Participants were unaware (blue) of 21.2% of errors and had a similar percent of no responses following *Stroop* (gray: 5.2%) and *repeat errors* (gray: 5.8%; *t*[*102*]=-0.7, *p*=0.5). <u>**B)**</u> When considering aware errors, RTs were *faster* on the first (356±19ms) and second post-error Go trials (364±11ms) relative to the pre-error Go trial (444±7ms; *F*[*2,170*]=14.4, *p*<0.001). In contrast, for unaware errors, RTs were *slower* on the first post-error Go trial (523±12ms) and did not differ for the second post-error trial (462±9ms) relative to pre-error RTs (448±9ms; *F*[*2, 170*]=15.1, *p*<0.001).

Also consistent with prior task implementations [49, 89-91], participants were aware of most errors (73.4±2.4%), were unaware of 21.2±2.3%, and failed to respond after 5.4±0.7% of errors (**Table 2, Fig. 1A**). To assess group differences in error awareness, we performed a 2(HIV) x 2(CB) x 2(AWARENESS: aware vs. unaware) ANOVA. While we detected an AWARENESS main effect (*F*[*1, 99*]=106.5, *p*<0.001) indicating that participants were aware of most errors, we did not observe any significant group-related main effects or interactions (*p*’s>0.9). Consistent with previous reports [49, 89, 90], participants failed to indicate awareness more often following repeat (28.6±2.8%) versus Stroop trials (15.5±2.1%; *t*[102]=7.7, *p*<0.001; **Fig. 1A**).

For awareness-related brain and behavioral assessments, only participants with at least two instances of aware and unaware errors were considered. Three participants (*n*=1 HIV+/CB+; *n*=1 HIV+/CB-; *n*=0 HIV-/CB+; *n*=1 HIV-/CB-) were excluded from further analyses as they indicated awareness of too few errors, suggesting that they did not understand the error awareness signaling procedure. Another 14 participants (*n*=8 HIV+/CB+; *n*=3 HIV+/CB-; *n*=1 HIV-/CB+; *n*=2 HIV-/CB-) were excluded as they had too few unaware trials (i.e., they detected all errors). This yielded a sample of 86 participants available for error awareness assessments.

To assess RT patterns following (un)aware errors, we performed a 2(HIV) x 2(CB) x 2(AWARENESS) x 3(TRIAL: pre-vs. 1-post-vs. 2-post-error) mixed-effects ANOVA. While no group-related effects were observed, an AWARENESS x TRIAL interaction was detected (*F*[*2, 81*]=53.1, *p*<0.001; **Fig. 1B**). Follow-up tests identified a main effect of TRIAL when considering RTs associated with both aware (*F*[*2,170*]=14.4, *p*<0.001) and unaware errors (*F*[*2,170*]=15.1, *p*<0.001). These main effects were driven by two distinct post-error (vs. pre-error) RT patterns. Specifically, RTs were faster on the Go trial immediately following (vs. preceding) an aware error (1-post-error: *t*[*85*]=4.2, *p*<0.001), yet RTs were slower following an unaware error (1-post-error: *t*[*85*]=-4.4, *p*<0.001). As RTs on the first post-error trial (i.e., the awareness trial) were confounded with the error signaling process, we also considered RTs on the subsequent Go trial (2-post-error). We again observed faster RTs (speeding-up) following aware errors (2-post-error: *t*[*85*]=9.5, *p*<0.001), but detected no such RT difference following unaware errors (*t*[*85*]=-1.8, *p*=0.07). This post-error speeding after aware, yet post-error slowing after unaware errors, is consistent with prior EAT findings [89]. We speculate that these RT patterns reflect an ‘eagerness’ to indicate error awareness following aware errors that is not present after unaware errors.

#### 3.3.1. Task effects: Cognitive control/failure-related brain activity

To characterize activity differences during cognitive control failures (errors) versus successes (inhibitions), we contrasted BOLD signal changes associated with NoGo-error [E] versus NoGo-correct trials [C] (**Fig. 2A**). Increased activity during error trials [E>C] was observed notably in the bilateral aI, dmPFC (**Fig. 2B**), thalamus, and a large cluster encompassing the left primary and supplementary motor areas. In contrast, during correct trials [C>E], increased activity was observed in bilateral dorsal medial parietal cortex (dmPC), the bilateral putamen extending into the nucleus accumbens, the bilateral hippocampi, and the occipital lobe. We then assessed relations between error-related brain activity and the number of NoGo-errors committed (ERROR COUNT), an objective laboratory-based measure of cognitive failures. Increased error-related activity in the left aI (*r*[101]=-0.3, *p*=0.02) and decreased error-related activity in the dmPC (*r*[*101*]=0.4, *p*<0.004; **Fig. 2C**) correlated with fewer errors. We also considered brain-behavior relationships focusing on CFQ total scores, a self-reported measure of real-world cognitive failures (**Fig. 2D**). HIV serostatus moderated the association between self-reported cognitive failures and error-related left aI (HIVxCFQ: *F*[*1,99*]=4.0, *p*=0.049) and dmPFC activity (*F*[*1,99*]=9.1, *p*=0.003). Follow-up examination of within-group conditional effects indicated that among HIV-participants *more* error-related activity in the left aI (*β*=-0.02, *p*=0.02) and dmPFC (*β*=-0.02, *p*=0.03) was linked with fewer self-reported cognitive failures. Different patterns were observed among PLWH such that error-related aI activity was not correlated with self-reported cognitive failures (*β*=0.003, *p*=0.7) and *less* dmPFC activity was related to fewer cognitive failures (*β*=0.01, *p*=0.04).

**Figure 2.**
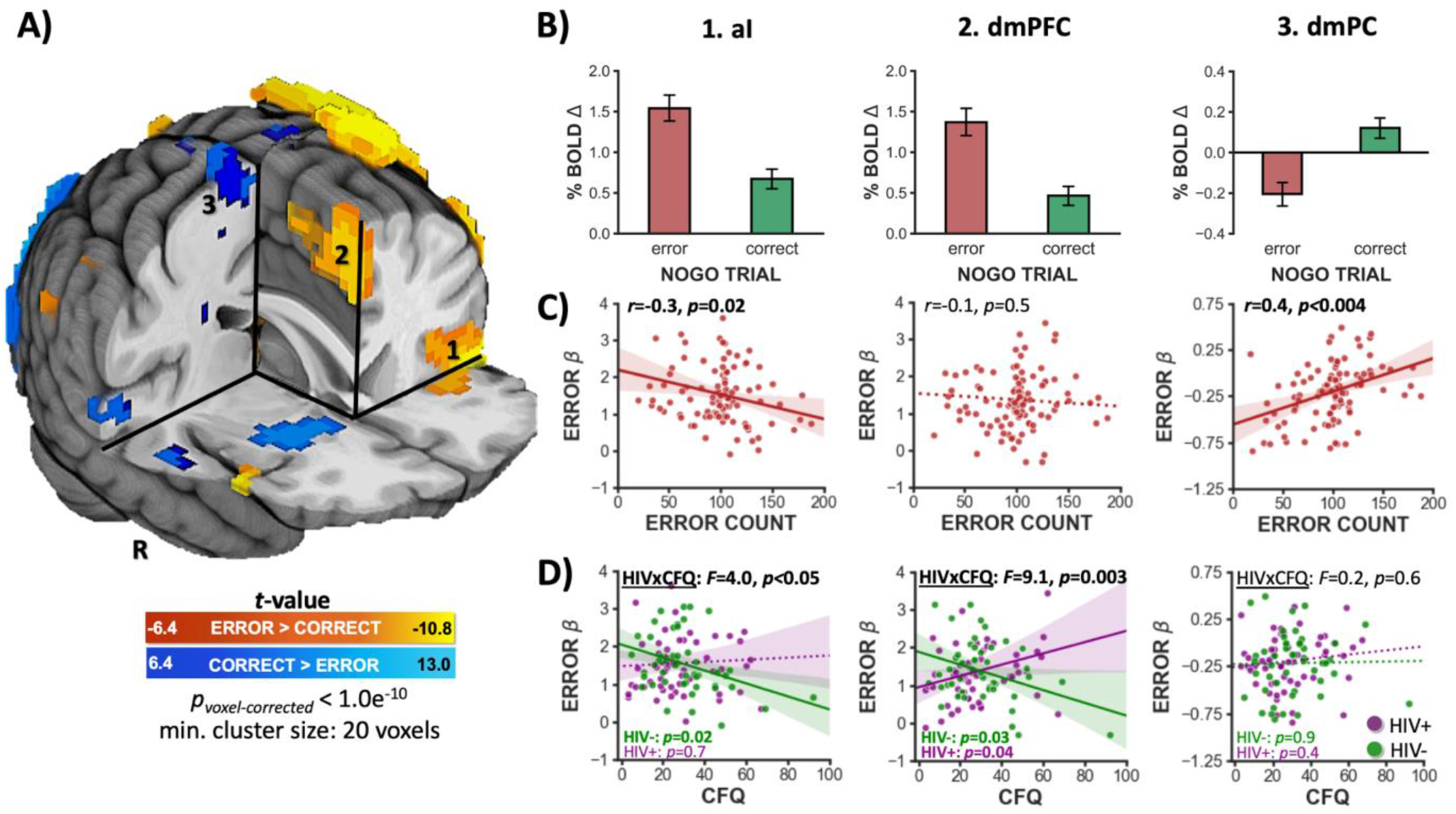
Whole-brain activity following error [E] and correct [C] NoGo trials. <u>**A)**</u> Across the sample of 103 participants demonstrating sufficient task engagement (i.e., responded to >50% of Go trials), cognitive control failures (errors) were associated with increased activity in the bilateral anterior insula (aI), dorsal medial prefrontal cortex (dmPFC), left thalamus, and left primary motor cortex extending into supplementary motor regions (warm colors, E>C). On the other hand, cognitive control successes (inhibitions) were associated with increased activity in the bilateral putamen and dorsal medial parietal cortex (dmPC; cold colors, C>E; *p*_*corrected*_<1.0e^- 10^). <u>**B)**</u> Mean percent BOLD signal change (*β*) values for NoGo-error and -correct trials from the (1) left aI, (2) dmPFC, and (3) right dmPC (numbering corresponds to that shown in panel A). <u>**C)**</u> Correlations between error-related *β* values from each ROI and an objective measure of cognitive failures. More error-related left aI activity (*p*_*Bonferronni-corrected*_=0.02) and more right (*p*_*Bonferronni-corrected*_<0.004) and left dmPC activity [*r*(*101*)=0.4, *p*_*Bonferronni-corrected*_<0.004, data not shown] correlated with fewer NoGo errors. **D)** Significant HIV x CFQ interactions were detected when considering error-related left aI (*p*=0.049) and dmPFC activity (*p*=0.003) such that more activity was linked with less self-reported cognitive failures among HIV-participants (green), but not among HIV+ participants (purple). See **Supplemental Table S7** for cluster coordinates.

#### 3.3.2. Task effects: Error awareness-related brain activity

To characterize activity linked with explicit error awareness, we contrasted BOLD signal changes associated with Aware [A] versus Unaware [U] NoGo-errors (**Fig. 3A**). Increased activity during aware error trials [A>U] was observed notably in the bilateral insulae, the bilateral putamen (**Fig. 3B**), the posterior thalamus, and in bilateral clusters extending into the dmPFC, and encompassing the primary and supplementary motor areas. In contrast, during unaware errors [U>A], we observed increased activity in a right mPFC cluster and reduced deactivation in the bilateral PCC (**Fig. 3B**). We then assessed relationships between awareness-related brain activity ([A-U] contrast values) and the percent of unaware errors (% UNWARE), an objective measure of explicit error awareness. Increased awareness-related brain activity in the right putamen was associated with reduced awareness behavioral performance, although this association failed to reach significance following multiple comparisons correction (*r*[*84*]=0.3, *p*=0.07; **Fig. 3C**). When considering brain-behavior relationships focusing on CFQ scores (**Fig. 3D**), HIV did not moderate the relationship between awareness-related brain activity and self-reported cognitive failures. Rather, we observed a main effect of CFQ in the left PCC (*F*[*1, 82*]=4.3, *p*=0.04) such that, across both PLWH and HIV-participants, more deactivation following aware (vs. unaware) errors correlated with fewer self-reported cognitive failures. We did not observe any relationships between CFQ and % UNAWARE (*p*’s>0.09). At the whole-brain level, *no group effects* were detected when considering the [A-U] contrast images. However, follow-up exploratory analyses assessing group effects on averaged NoGo-error-aware [A] and NoGo-error-unaware [U] *β* coefficients within selected ROIs are reported in the supplemental material (**Supplemental Figure S3**).

**Figure 3.**
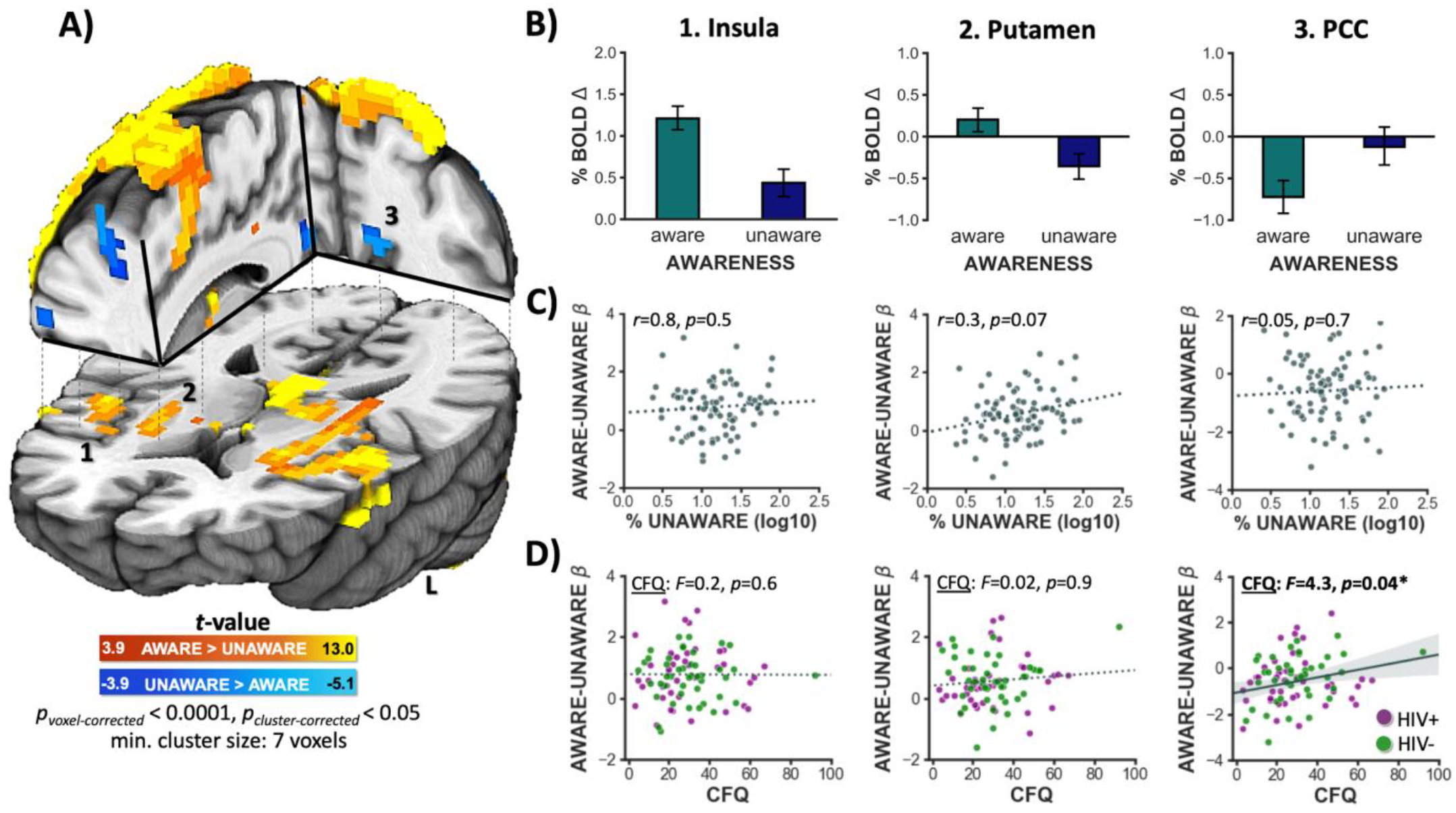
Whole-brain activity following aware [A] and unaware [U] errors. <u>**A)**</u> Across the sample of 86 participants committing a sufficient number of aware and unaware errors, explicit error awareness was associated with increased activity in the bilateral insula, bilateral putamen, posterior thalamus, and bilateral primary motor cortex extending into supplementary motor regions (warm colors: A>U, *p*_*corrected*_<0.05). On the other hand, unaware error trials were associated with increased activity (i.e., *less deactivation*) of the bilateral posterior cingulate cortex (PCC) (cold colors: U>A). <u>**B)**</u> Mean percent BOLD signal change (*β*) values for aware and unaware error trials for the (1) right insula, (2) right putamen, and (3) left PCC. <u>**C**)</u> Correlations between the percent of unaware errors and [A-U] contrast values from each ROI. More aware (vs. unaware) error-related brain activity in the right putamen correlated with a higher percentage of unaware errors, but this association did not survive multiple comparison corrections (*p*_*Bonferronni-corrected*_ =0.07). <u>**D**)</u> A main effect of CFQ total scores on left PCC (*p*=0.04) awareness-related activity was detected such that across all participants, more PCC deactivation following aware (vs. unaware) errors was linked with fewer self-reported cognitive failures. See **Supplemental Table S8** for cluster coordinates.

### 3.4. Group effects: Cognitive control/failure-related brain activity

To delineate functional brain alterations as a function of HIV and CB, we assessed [E-C] contrast images in a whole-brain ANOVA. While we did not detect any clusters displaying HIV x CB or CB main effects, we did observe HIV main effects in the PCC and mPFC (**Fig. 4A**). Graphical assessment of *β* values extracted from these ROIs indicated that HIV+ (vs. HIV-) participants displayed less regional deactivation following errors, but not correct trials (**Fig 4B**). To further characterize PCC and mPFC activity dynamics across EAT trials, we conducted qualitative inspection and follow-up assessments when considering *β* coefficients for the three Go trials both preceding (1-pre, 2-pre, and 3-pre) and succeeding a NoGo-error (1-post [awareness trial], 2-post, 3-post) (**Fig. 4C**). Both the PCC (*F*[*5,505*]=4.1, *p*=0.03) and mPFC (*F*[*5,505*]=6.6, *p*=0.002) displayed a significant HIVxTRIAL interaction such that HIV-participants displayed deactivation on the Go trials immediately preceding NoGo-errors (1-pre), displayed the most deactivation on error trials, and this deactivation persisted into the next trial (1-post/awareness trial). In contrast, PLWH maintained relatively stable PCC and mPFC activity across trial types. As the two groups displayed large activity differences on error trials, we assessed the relation between error-related PCC and mPFC deactivations and cognitive control performance (i.e., NoGo-error count; **Fig. 4D**). Among HIV-participants, more error-related PCC deactivation was linked with fewer errors (*p*=0.048), whereas among PLWH the opposite pattern was observed such that more PCC deactivation was linked with more errors (*p*=0.049).

**Figure 4.**
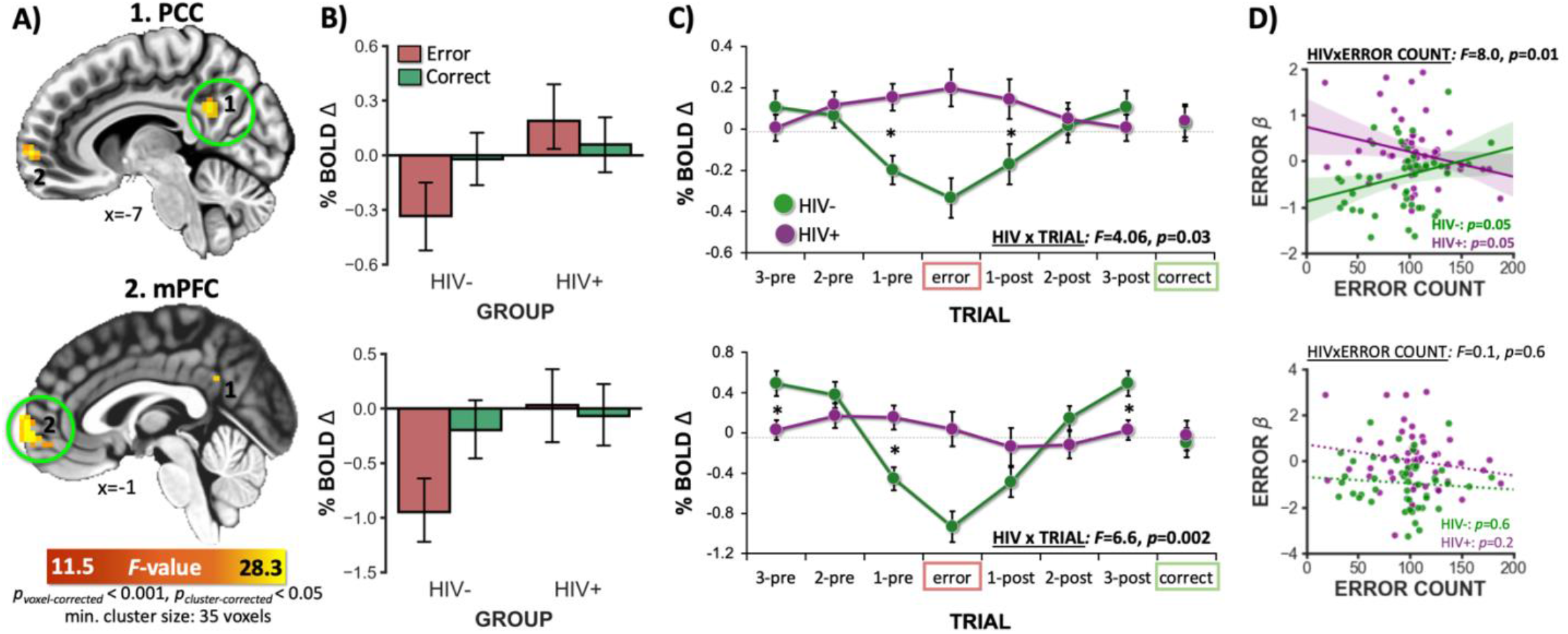
HIV+ vs. HIV-group differences in error-related brain activity. **A)** HIV main effects were observed in the posterior cingulate cortex (PCC, 39 voxels) and medial prefrontal cortex (mPFC, 36 voxels; whole-brain HIV x CB ANOVA, *p*_*corrected*_<0.05) when considering NoGo correct vs. error [C-E] contrast values. **B**) Qualitative inspection of BOLD signal change within identified ROIs indicated that, during errors (red bars), the HIV- (vs. HIV+) group displayed more deactivation in the PCC (1) and mPFC (2). **C**) *β* coefficients during NoGo-errors, and the three Go trials both preceding (1-pre, 2-pre, and 3-pre) and following a NoGo-error (1-post [awareness trial], 2-post, 3-post) plotted sequentially with respect to NoGo-correct *β* coefficients for comparison. We detected HIV x TRIAL interactions for the PCC (*p*_*Bonferronni-corrected*_=0.03) and mPFC ROIs (*p*_*Bonferronni-corrected*_=0.002; NoGo-error and NoGo-correct *β* coefficients were not included in this statistical test to avoid circular analyses). Qualitative inspection indicated that both PCC and mPFC activity among HIV-participants displayed a deactivation the trial before an error, showed the most deactivation following errors, and that this deactivation persisted into the next trial. In contrast, PLWH showed relatively little modulation of ROI activity across the various trials (*’s indicate significant difference between HIV groups in follow-up *t*-tests). **D**) Significant HIV x ERROR COUNT interactions on error-related PCC [E] *β* coefficients (*F*[*1,99*]=6.0, *p*_*Bonferronni-corrected*_ =0.01). Among HIV-participants, more PCC deactivation following errors correlated with fewer NoGo errors, whereas among PLWH the opposite pattern was observed. See **Supplemental Table S9** for cluster coordinates.

### 3.5. Real-world implications: Error-related PCC deactivations correlated with medication management ability

When controlling for biological sex and lifetime CB use, PLWH displayed poorer MMT performance (10.8±0.5) relative to HIV-participants (11.9±0.4; *F*[*1,97*]=8.0, *p*=0.006). However, self-reported cognitive failures (CFQ total scores) did not differ between HIV groups (*F*[*1,97*]=0.4, *p*=0.5) and no correlation between MMT performance and CFQ scores was detected (*r*[*101*]=-0.04, *p*=0.7). We then considered brain-behavior relationships focusing on MMT performance. We observed that error-related PCC deactivations were associated with MMT performance (*F*[*1,99*]=11.4, *p*=0.001; **Fig. 5A**) such that enhanced deactivation was linked with better medication management across both HIV groups (HIVxMMT: *F*[*1,99*]=0.2, *p*=0.9). Next, we formally tested the relationship between these variables in a mediation model. PCC deactivation (M) mediated the effect of HIV serostatus (X) on medication management (Y; CI95% = −0.05, −0.06; **Fig. 5B**) such that PLWH displayed reduced error-related PCC deactivation (*p*<0.001) which, in turn, was associated with poorer MMT performance (*p*<0.001). Pathways remained unchanged when including sex and lifetime CB use as covariates. Given limitations of the cross-sectional and observational nature of our study design, we do not assume causality between these variables [106, 107]. We did not detect any significant relationships between error-related ROI activity and self-reported cognitive failures (*p*’s>0.3; **Fig 5C**).

**Figure 5.**
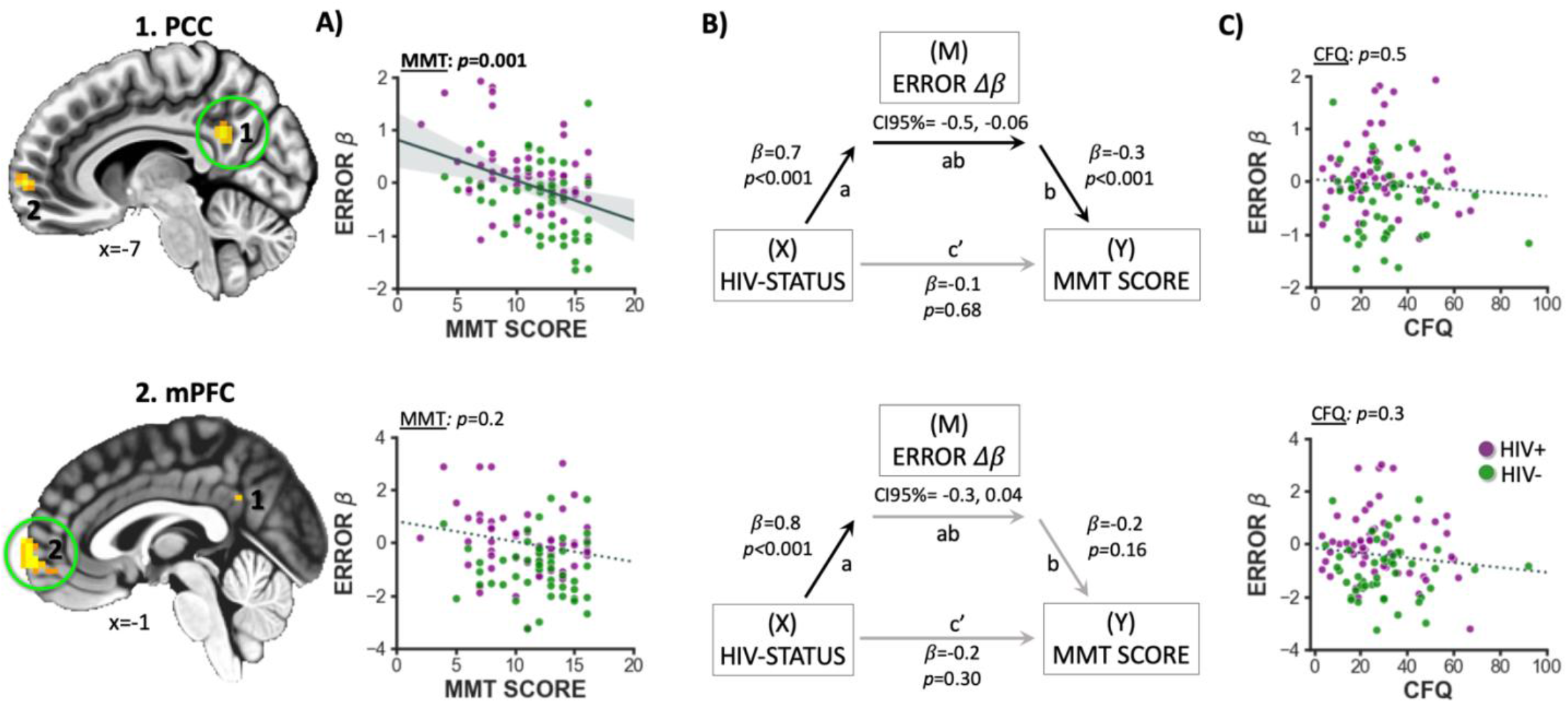
Relations between error-related brain activity showing HIV main effects and medication management performance. **A**) Greater error-related PCC deactivation was linked with better MMT performance (ANCOVA, MMT main effect: *p*=0.001) among both HIV+ (*p*=0.01) and HIV-(*p*=0.03) individuals. **B**) Error-related PCC activity (M) mediated (CI95%=0.01, 0.14) the relationship between HIV-status (X) and MMT scores (Y) such that, the HIV+ group had reduced PCC deactivation which was associated with poorer MMT performance. The mediation model was not significant for mPFC error activity (CI95%= −0.01, 0.10). **C**) Error-related PCC and mPFC activity was not correlated with self-reported cognitive failures (CFQ total scores, *p*’s>0.3).

### 3.6. Impact of chronic CB use on brain and behavior

While we did not detect differences among CB using groups when considering error-related PCC or mPFC activity (*p*’s>0.8), we observed that lifetime CB use impacted the magnitude of mPFC deactivations among HIV-participants, but not among PLWH. Specifically, when controlling for past month nicotine use, we observed a significant HIV x LIFETIME AMOUNT interaction when considering mPFC activity (*F*[*1,54*]=4.7, *p*=0.04; **Fig. 6A**). Follow-up within-group conditional effects indicated that among HIV-CB users, more use was linked with less error-related mPFC deactivation (*β*=0.7, *t*[*24*]=2.0, *p*=0.06). Whereas, among CB using PLWH, mPFC activity was not correlated with lifetime use (*β*=-0.2, *t*[*28*]=-0.7, *p*=0.5). We did not observe any significant associations of LIFETIME AMOUNT on error-related PCC activity. Similar effects were observed when assessing times using CB over the lifetime (**Supplemental Figure S2**). Interestingly, while CB users did not display worse medication management performance relative to non-users (*F*[*1,99*]=0.8, *p*=0.4), across all CB users, we observed a relationship between LIFETIME AMOUNT and MMT scores (**Fig. 6B**) such that, more use was linked with poorer MMT performance (*F*[*1,50*]=8.5, *p*=0.005). No relationship between CB use and self-reported cognitive failures was observed (CFQ; *p’s*>0.8; **Fig. 6C**).

**Figure 6.**
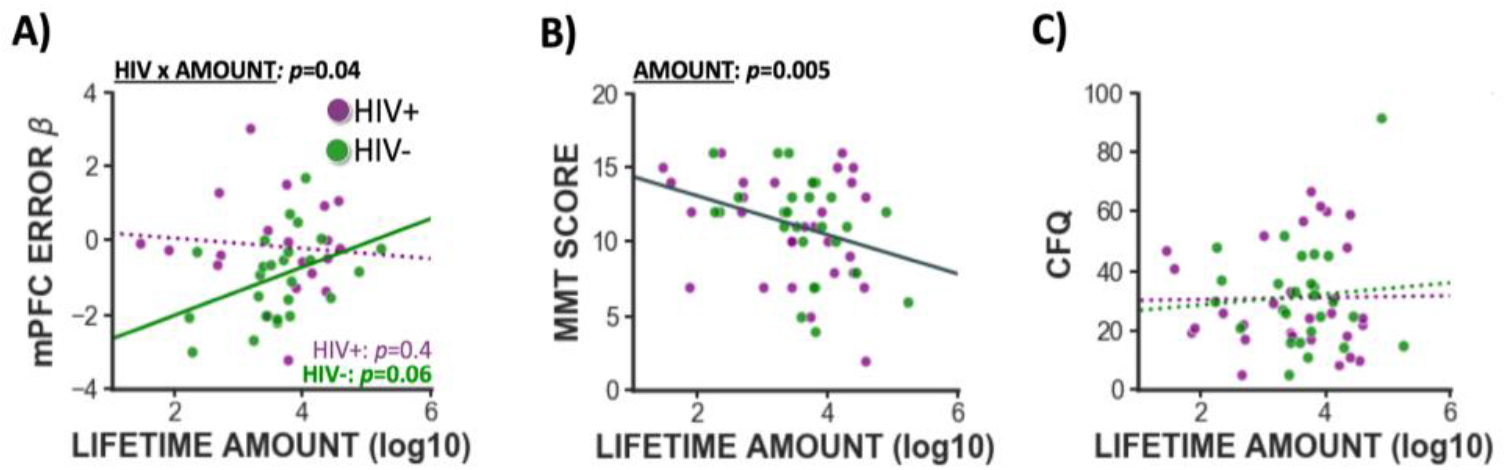
Impact of CB use on brain and behavior. **A**) Among HIV-controls, reduced error-related mPFC deactivations were linked with more lifetime CB use (although the within-group conditional effect did not reach significance; *p*=0.06) whereas, among PLWH, error-related mPFC activity was not associated with use. **B**) When controlling for past month nicotine use, more lifetime CB use was associated with poorer MMT performance (*p*=0.005) similarly for both HIV+ and HIV-groups (HIV x AMOUNT: *p*>0.4) **C**) Lifetime CB use was not associated with self-reported cognitive failures (CFQ total score; *p’s*>0.8).

## DISCUSSION

HIV is characterized by a progressive neurocognitive decline that can impact everyday functions, including those critical for ongoing disease management. Emerging evidence indicates that PLWH also exhibit a lack of insight into their cognitive deficits which may manifest in the laboratory as diminished error recognition [18-21]. To delineate altered error-related brain activity among PLWH, we employed a modified Go/NoGo task where participants indicated awareness of their errors. We observed that error-related aI activity was linked with fewer objectively measured commission errors across all participants and fewer subjectively reported cognitive failures among HIV-participants. Regarding error awareness, more insula *activation* and more PCC *deactivation* was linked with aware (vs. unaware) errors, and such PCC deactivation correlated with fewer self-reported cognitive failures across all participants. Regarding group effects, PLWH lacked error-related deactivation in two DMN regions (i.e., PCC, mPFC), deactivations which were observed among HIV-participants and were associated with fewer commission errors. Importantly, we also documented clinically-relevant implications for such altered PCC responsivity, such that reduced deactivations were linked with poorer medication management abilities across all participants. Regarding CB-related effects, lifetime use was associated with reduced error-related mPFC deactivation among CB using HIV-participants as well as poorer medication management abilities across all CB users. Taken together, these results demonstrate that insufficient DMN suppression, linked with HIV infection and chronic CB use among HIV-participants, has real-world consequences for medication management abilities.

### 4.1. EAT behavioral outcomes replication

Increasing confidence in our neuroimaging outcomes, we largely replicated previously reported behavioral effects within the EAT. First, error and error awareness rates observed across our sample were similar to those previously documented among nondrug using [89], CB using [49], and cocaine using participants [91]. Second, our participants demonstrated lower accuracy, yet higher awareness of errors under the NoGo: Stroop (vs. repeat) condition which is consistent with prior studies (e.g., [49, 90]) and the interpretation that the NoGo: Stroop rule was more difficult to monitor [89]. Third, we replicated previously reported RT reductions on Go trials following aware errors, yet RT slowing following unaware errors [49, 89, 90]. Such RT differences bolster the argument that these two error types are qualitatively distinct and differentially processed by the brain. Despite these task-related behavioral replications, we did not observe anticipated group differences when considering number of errors, percent of aware errors, or RT outcomes. Such null effects were initially surprising given that HIV infection has been linked with reduced processing speed [109] and slower RTs in a Stroop task [110] and that CB use has been linked with reduced error awareness [49]. We speculate that the lack of HIV-related effects observed herein, particularly regarding error rate and RT, may be related to the dynamically adapting EAT difficulty manipulation.

### 4.2. EAT neuroimaging outcomes replication

Utilizing a contrast of NoGo correct versus error [C-E] trials, we probed brain activity differentially modulated during cognitive control success and failures. Consistent with meta-analytic outcomes compiling neuroimaging results from cognitively demanding Go/NoGo tasks [57, 111], we observed increased *cognitive control-related* brain activity on correct trials in the dmPC, bilateral putamen, hippocampus, and occipital areas and, conversely, increased *error-related* activity in the aI, dmPFC, as well as primary and supplementary motor cortex. Increased aI and dmPFC activity has been consistently linked with error processing [112-115] and, more generally, the monitoring of external cues indicating the need for increased cognitive control deployment to achieve behavioral goals [92, 116]. As such, error-related aI and dmPFC responsivity may facilitate behavioral adaptations to avoid future negative outcomes [116]. Supporting this notion, we observed that increased error-related aI activity was correlated with better cognitive control performance (i.e., fewer NoGo errors) across all participants and less self-reported everyday cognitive failures among HIV-participants. In contrast, among PLWH, a similar relationship between aI activity and self-reported cognitive failures was not observed, which we suggest is reflective of a lack of insight into cognitive alterations linked with HIV infection.

Utilizing a contrast of NoGo aware versus unaware [A-U] errors, we probed brain activity related to explicit error awareness and observed increased activation in the bilateral insulae, putamen, occipital lobe, and primary and supplementary motor areas. Our findings replicate prior work implicating the insula in error awareness [49]. Moving beyond error awareness, the insula is theorized to play a critical role in interoceptive, emotional, and other forms of subjective awareness [117] which may have particular significance for insight into cognitive and affective alterations across neuropsychiatric conditions including addiction [55]. Notably, we also observed greater bilateral PCC deactivations during aware (vs. unaware) errors which correlated with fewer self-reported cognitive failures across all participants and, unlike insula responsivity, has not been as widely emphasized in previous EAT neuroimaging studies [49]. While, other work has linked reduced insular volume to unawareness of memory loss among patients diagnosed with Alzheimer’s Disease [118] and reduced insight among patients diagnosed with Schizophrenia [119], our findings implicating PCC deactivations with both objective error awareness and subjective reports of cognitive failures, indicate that the PCC may be critically important for insight into cognitive alterations. Our PCC findings are also consistent with a key role for this region in awareness [120, 121] and other work linking dynamic DMN activity with attention [122], stimulus detection [64, 66], and intermittent cognitive failures [122-124],

### 4.3. HIV-associated reductions in error-related DMN suppression

Unlike HIV-participants, PLWH displayed a lack of error-related deactivation in two DMN regions (i.e., PCC, mPFC). Whereas HIV-participants displayed robust PCC and mPFC deactivations following NoGo errors (and to a certain extent, on the trials immediately preceding and succeeding those errors), such DMN regional activity among PLWH remained relatively stable across trial types. Such deactivations play a causal role in task-vigilance [125] such that insufficient DMN suppression often precedes and is predictive of intermittent attentional lapses [122-124, 126]. Primate electrophysiology work has also shown that PCC neurons become deactivated following errors and, importantly, that such deactivation is necessary for new learning [127]. Taken together, such observations suggest that insufficient error-related DMN suppression may contribute to poorer task performance and the disruption of behavioral adaptation [69, 127]. Such DMN suppression may reflect the reallocation of limited cognitive resources from task-irrelevant mental operations toward external task demands [69]. EAT errors are particularly salient task events, indicating that the participant must now execute an additional task behavior (i.e., press button-2 on the next Go trial [Awareness-trial] instead of the usual button-1). As such, error-related DMN suppression may be advantageous and reflect the re-deployment of attentional resources toward the task in preparation for the subsequent Awareness-trial and the avoidance of future errors. Indeed, greater error-related PCC deactivations were linked with fewer errors among HIV-participants. We speculate that a lack of error-related DMN suppression among PLWH is indicative of a reduced ability to disengage task irrelevant mental operations and re-deploy attentional resources toward the task. Whereas altered task-related DMN suppression has been linked with various neuropsychiatric conditions [53, 68-76], one differentiating aspect of our study is linking this cognitive neuroscience mechanism to HIV-associated neurobiological alterations and clinically-relevant implications.

### 4.4. Real-world implications of HIV-associated brain alterations

Speaking to clinically-relevant implications, across all participants, we observed that reduced error-related PCC deactivation correlated with poorer performance on a behavioral measure of medication management abilities. Accumulating evidence links poorer HIV medication management with worse neurocognitive outcomes, particularly executive dysfunction [85, 86, 128-130]. Although antiretroviral therapies significantly improve clinical outcomes and prolong life for PLWH [23, 24], strict medication adherence is crucial for success [24-27] and adherence rates remain unacceptably low (40-67%) [33]. Our results indicate that diminished error-related PCC deactivation among PLWH may contribute, in part, to these low adherence rates. We speculate that insufficient error-related DMN suppression may reflect an individual’s reduced ability to disengage distracting thoughts and to maintain attentional focus on a complex task. Providing some support for this perspective, we observed that the magnitude of error-related PCC deactivation mediated the effect of HIV serostatus on medication management abilities. We speculate that interventions facilitating DMN suppression (e.g., mindfulness-based practices, working-memory training, [131, 132]), may be beneficial for PLWH and the challenges they face. Noteworthy, self-reported cognitive failures were not significantly associated with medication management ability potentially suggesting a lack of self-awareness of one’s cognitive abilities which has been previously highlighted in the HIV literature [133, 134].

### 4.5. Impact of chronic CB use on brain and behavior

We found that lifetime amount of CB used was correlated with reduced error-related mPFC deactivation among HIV-controls and reduced medication management abilities across all participants. Specifically, more CB use by HIV-participants was linked with reduced mPFC deactivations such that, at higher levels of use, HIV-participants’ brain responsivity approached that of PLWH. Our results are consistent with other EAT studies that have documented reduced mPFC deactivations following NoGo errors among drug users (i.e., ecstasy, cannabis), in the absence of behavioral performance deficits [135]. In addition, other studies have linked chronic CB use with increased mPFC activity across a variety of cognitive and emotional tasks [136]. While we did not observe group differences (CB+ vs. CB-) in medication management abilities or brain activity, we did find that, among CB users, more lifetime use was linked to poorer medication management abilities. Given the complex effects of CB use duration [43], heaviness [40, 42], and age of regular use onset [44] on cognitive alterations among PLWH, our results may suggest that CB’s impact on the brain and behavior is dependent on use history. However, within the range present in our sample of PLWH, amount of lifetime use was not associated with further reductions in error-related mPFC deactivation, suggesting that the impact of CB use on medication management abilities, may be related to another neurobiological mechanism.

### 4.6. Limitations

Our experimental design allowed us to consider the separate and combined effects of HIV and CB on cognitive control and error-related brain activity. Yet, our results should be considered in light of methodical limitations. First, given the design’s cross-sectional nature, we cannot determine whether group differences in brain or behavior are caused by HIV and/or drug use or whether they represent other preexisting social, environmental, genetic, or personality risk factors that may predispose one to contracting HIV or using CB. Large-scale, longitudinal research is needed to disentangle the antecedents and consequences of both HIV and CB use. Second, while we collected a broad range of demographic, health, and cognitive-behavioral data from participants, additional variables that may have been of interest were not available. Specifically, socioeconomic status (SES) is known to exert a profound impact on a wide range of health and quality of life outcomes with many indirect downstream consequences. As such, the impact of SES on HIV and CB effects on the brain should be considered in future work. Third, we were unable to characterize the impact of CB use onset before versus after HIV seroconversion due to lack of power. Fourth, results have been mixed regarding the MMT-R’s ability to map to self-reported medication adherence [87]. This is challenging as self-report assessments of medication adherence may not be accurate particularly when considering potential lack of insight [18, 137]. We suggest that this validated, objective behavioral measure of medication management ability, provides a useful proxy for an individuals’ capacity to adhere to complex medication regimens. Nonetheless, future research with improved operationalizations of actual medication adherence may want to examine other factors beyond medication management ability (e.g., social, environmental, financial) influencing adherence outcomes. Finally, although often implicit in mediation models, we do not assume causality between variables included given the cross-sectional and observational nature of our study design [106, 107]. Longitudinal designs and/or those implementing experimental manipulations are better suited for establishing causal relationships in mediation models [107, 138-140].

### 4.7. Conclusions

Our results demonstrate insufficient error-related DMN suppression linked with HIV infection, as well as chronic CB use among HIV-participants, and associated with clinically-relevant consequences for medication management behaviors. Delineating this cognitive neuroscience mechanism may provide heuristic value for strategies to improve medication adherence. As insufficient DMN suppression appears to be a common endophenotype across various neuropsychiatric conditions, our results further highlight the importance and ubiquity of this cognitive neuroscience perspective. Given robust evidence that DMN suppression is linked with attention toward external stimuli, we posit that certain HIV and CB-associated neurocognitive alterations may stem from a reduced ability to disengage task irreverent mental operations that ultimately hinder cognitive control, error processing, and behavioral adaptation.

## Data Availability

The authors have released all code associated with this manuscript. The code and tabular data are available on GitHub (https://github.com/Flanneryg3/HIVCB_ProjectCode), and unthresholded brain maps are available on NeuroVault (https://neurovault.org/collections/9337/).

https://neurovault.org/collections/9337/

## ACKNOWLDGEMENTS

The authors have no conflicts to declare. Primary support for this project was provided by the National Institutes of Health (NIH) [K01DA037819] (JSF, MTS), and [R01DA033156] (RG), and the Florida International University (FIU) Graduate School Dissertation Year Fellowship (JSF). Contributions from authors were also provided with support from NIH [U54MD012393] (JSF, MTS), [R01DA041353] (MTS, ARL, MCR, RP), and National Science Foundation (NSF) [1631325] (ARL, MCR, TS). We thank the FIU Instructional & Research Computing Center (IRCC, http://ircc.fiu.edu) for providing access to the HPC computing resources that contributed to the generation of the research results reported herein.

## Notes

### Competing Interest Statement

The authors have declared no competing interest.

### Author Declarations

Florida International University's Institutional Review Board

## REFERENCES

1. Campbell, J.H., et al., Anti-alpha4 antibody treatment blocks virus traffic to the brain and gut early, and stabilizes CNS injury late in infection. PLoS Pathog, 2014. 10(12): p. e1004533.10.1371/journal.ppat.1004533

2. Hong, S. and W.A. Banks, Role of the immune system in HIV-associated neuroinflammation and neurocognitive implications. Brain, behavior, and immunity,, 2015. 45: p. 1–12

3. Anthony, I.C., et al., Influence of HAART on HIV-related CNS disease and neuroinflammation. Journal of Neuropathology & Experimental Neurology, 2005. 64(6): p. 529–536.https://doi.org/10.1093/jnen/64.6.529

4. Melrose, R.J., et al., Compromised fronto-striatal functioning in HIV: an fMRI investigation of semantic event sequencing. Behav Brain Res, 2008. 188(2): p. 337–47.10.1016/j.bbr.2007.11.021

5. Plessis, S.D., et al., HIV infection and the fronto-striatal system: a systematic review and meta-analysis of fMRI studies. AIDS, 2014. 28(6): p. 803–11.10.1097/QAD.0000000000000151

6. Holt, J.L., S.D. Kraft-Terry, and L. Chang, Neuroimaging studies of the aging HIV-1-infected brain. J Neurovirol, 2012. 18(4): p. 291–302.10.1007/s13365-012-0114-1

7. Maki, P.M., et al., Impairments in memory and hippocampal function in HIV-positive vs HIV-negative women: a preliminary study. Neurology, 2009. 72(19): p. 1661–1668. https://doi.org/10.1212/WNL.0b013e3181a55f65

8. Castelo, J.M.B., et al., Altered hippocampal-prefrontal activation in HIV patients during episodic memory encoding. Neurology, 2006. 66(11): p. 1688–1695

9. Kuper, M., et al., Structural gray and white matter changes in patients with HIV. J Neurol, 2011. 258(6): p. 1066–75.10.1007/s00415-010-5883-y

10. Abdullaev, Y., et al., Functional MRI evidence for inefficient attentional control in adolescent chronic cannabis abuse. Behavioural brain research, 2010. 215(1): p. 45–57

11. Burggren, A.C., et al., Cannabis effects on brain structure, function, and cognition: considerations for medical uses of cannabis and its derivatives. Am J Drug Alcohol Abuse, 2019: p. 1–17.10.1080/00952990.2019.1634086

12. Kroon, E., et al., Heavy cannabis use, dependence and the brain: a clinical perspective. Addiction, 2019.10.1111/add.14776

13. Behan, B., et al., Response inhibition and elevated parietal-cerebellar correlations in chronic adolescent cannabis users. Neuropharmacology, 2014. 84: p. 131–137

14. Dawes, S., et al., Variable patterns of neuropsychological performance in HIV-1 infection. J Clin Exp Neuropsychol, 2008. 30(6): p. 613–26.10.1080/13803390701565225

15. Woods, S.P., et al., Cognitive neuropsychology of HIV-associated neurocognitive disorders. Neuropsychol Rev, 2009. 19(2): p. 152–68.10.1007/s11065-009-9102-5

16. Heaton, R., et al., HIV-associated neurocognitive disorders persist in the era of potent antiretroviral therapy: CHARTER Study. Neurology,, 2010. 75(23): p. 2087–2096

17. Mothobi, N.Z. and B.J. Brew, Neurocognitive dysfunction in the highly active antiretroviral therapy era. Curr Opin Infect Dis, 2012. 25(1): p. 4–9.10.1097/QCO.0b013e32834ef586

18. Hinkin, C., et al., Actual versus self-reported cognitive dysfunction in HIV-1 infection: memory-metamemory dissociations. Journal of Clinical and Experimental Neuropsychology, 1996. 18(3): p. 431–443

19. Van Gorp, W., et al., Metacognition in HIV-1 seropositive asymptomatic individuals: self-ratings versus objective neuropsychological performance. Journal of Clinical and Experimental Neuropsychology, 1991. 13(5): p. 812–819

20. Bassel, C., et al., Working memory performance predicts subjective cognitive complaints in HIV infection. Neuropsychology, 2002. 16(3): p. 400–10.10.1037//0894-4105.16.3.400

21. Vance, D., K. Farr, and S. T., Assessing the clinical value of cognitive appraisal in adults aging with HIV. Journal of Gerontological Nursing, 2008. 34(1): p. 36–41

22. Joska, J.A., et al., Does highly active antiretroviral therapy improve neurocognitive function? A systematic review. J Neurovirol, 2010. 16(2): p. 101–14.10.3109/13550281003682513

23. Murphy, E.L., et al., Highly active antiretroviral therapy decreases mortality and morbidity in patients with advanced HIV disease. Annals of internal medicine, 2001. 135(1): p. 17–26

24. Gauchet, A., T. Cyril, and F. Gustave, Psychosocial predictors of medication adherence among persons living with HIV. International Journal of Behavioral Medicine, 2007. 14(3): p. 141–150

25. Van Vaerenbergh, K., et al., A combination of poor adherence and a low baseline susceptibility score is highly predictive for HAART failure. Antiviral Chemistry and Chemotherapy, 2002. 13(4): p. 231–240

26. Paterson, D.L., et al., Adherence to protease inhibitor therapy and outcomes in patients with HIV infection. Annals of internal medicine, 2000. 133(1): p. 21–30

27. Bangsberg, D.R., Monitoring adherence to HIV antiretroviral therapy in routine clinical practice: The past, the present, and the future. AIDS Behav, 2006. 10(3): p. 249–51.10.1007/s10461-006-9121-7

28. Blaschke, T.F., Noncompliance and resistance to protease inhibitors, in Conference on Retroviruses and Opportunistic Infections. 1997.

29. Condra, J.H., Schleif, W., Blahy, O., Gabryelski, L. J. Graham, et al., In vivo emergence of HIV-1 variants resistant to multiple protease inhibitors. Nature, 1995. 374: p. 569–571

30. Deeks, S.G., Smith, M., Holodney, M., and Kahn, J., HIV-1 protease inhibitors: A review for clinicians. Journal of the American Medical Association, 1997. 277: p. 145–153

31. Albert S., M., et al., An Observed Performance Test of Medication Management Ability in HIV: Relation to Neuropsychological Status and Medication Adherence Outcomes. AIDS and Behavior, 1999. 3(2)

32. Albert, S.M., et al., Medication management skill in HIV: I. Evidence for adaptation of medication management strategies in people with cognitive impairment. II. Evidence for a pervasive lay model of medication efficacy.. AIDS and Behavior, 2003. 7(3): p. 329–338

33. Gao, X. and D.P. Nau, Congruence of three self-report measures of medication adherence among HIV patients. Annals of Pharmacotherapy, 2000. 34(10): p. 1117–1122

34. D’Souza, G., et al., Medicinal and recreational marijuana use among HIV-infected women in the Women’s Interagency HIV Study (WIHS) cohort, 1994-2010. J Acquir Immune Defic Syndr, 2012. 61(5): p. 618–26.10.1097/QAI.0b013e318273ab3a

35. Pacek, L.R., et al., Frequency of Cannabis Use and Medical Cannabis Use Among Persons Living With HIV in the United States: Findings From a Nationally Representative Sample. AIDS Educ Prev, 2018. 30(2): p. 169–181.10.1521/aeap.2018.30.2.169

36. Rizzo, M.D., et al., HIV-infected cannabis users have lower circulating CD16+ monocytes and IFN-gamma-inducible protein 10 levels compared with nonusing HIV patients. AIDS, 2018. 32(4): p. 419–429.10.1097/QAD.0000000000001704

37. Montgomery, L., et al., The Association Between Marijuana Use and HIV Continuum of Care Outcomes: a Systematic Review. Curr HIV/AIDS Rep, 2019. 16(1): p. 17–28.10.1007/s11904-019-00422-z

38. Manuzak, J.A., et al., Heavy Cannabis Use Associated With Reduction in Activated and Inflammatory Immune Cell Frequencies in Antiretroviral Therapy-Treated Human Immunodeficiency Virus-Infected Individuals. Clin Infect Dis, 2018. 66(12): p. 1872–1882.10.1093/cid/cix1116

39. Cristiani, S., N. Pukay-Martin, and R. Bornstein, Marijuana use and cognitive function in HIV-infected people. The Journal of neuropsychiatry and clinical neurosciences, 2004. 16(3): p. 330–335

40. Thames, A.D., et al., Combined effects of HIV and marijuana use on neurocognitive functioning and immune status. AIDS Care, 2016. 28(5): p. 628–32.10.1080/09540121.2015.1124983

41. Towe, S.L., et al., Reciprocal Influences of HIV and Cannabinoids on the Brain and Cognitive Function. J Neuroimmune Pharmacol, 2020.10.1007/s11481-020-09921-y

42. Thames, A.D., et al., Marijuana effects on changes in brain structure and cognitive function among HIV+ and HIV-adults. Drug Alcohol Depend, 2017. 170: p. 120–127.10.1016/j.drugalcdep.2016.11.007

43. Okafor, C.N., et al., Association of Marijuana Use with Changes in Cognitive Processing Speed and Flexibility for 17 Years in HIV-Seropositive and HIV-Seronegative Men. Subst Use Misuse, 2019. 54(4): p. 525–537.10.1080/10826084.2018.1495736

44. Skalski, L.M., et al., Memory Impairment in HIV-Infected Individuals with Early and Late Initiation of Regular Marijuana Use. AIDS Behav, 2018. 22(5): p. 1596–1605.10.1007/s10461-017-1898-z

45. Bonn-Miller, M.O., et al., Cannabis use and HIV antiretroviral therapy adherence and HIV-related symptoms. J Behav Med, 2014. 37(1): p. 1–10.10.1007/s10865-012-9458-5

46. Vidot, D.C., B. Lerner, and R. Gonzalez, Cannabis Use, Medication Management and Adherence Among Persons Living with HIV. AIDS Behav, 2017. 21(7): p. 2005–2013.10.1007/s10461-017-1782-x

47. Ernst, T., et al., Abnormal brain activation on functional MRI in cognitively asymptomatic HIV patients. Neurology,, 2002. 59(9): p. 1343–1349

48. Zhou, Y., et al., Motor-related brain abnormalities in HIV-infected patients: a multimodal MRI study. Neuroradiology, 2017. 59(11): p. 1133–1142.10.1007/s00234-017-1912-1

49. Hester, R., L. Nestor, and H. Garavan, Impaired error awareness and anterior cingulate cortex hypoactivity in chronic cannabis users. Neuropsychopharmacology, 2009. 34(11): p. 2450

50. Meade, C.S., et al., Synergistic effects of marijuana abuse and HIV infection on neural activation during a cognitive interference task. Addict Biol, 2018.10.1111/adb.12678

51. Harsay, H.A., et al., Error awareness and salience processing in the oddball task: shared neural mechanisms. Front Hum Neurosci, 2012. 6: p. 246.10.3389/fnhum.2012.00246

52. Ullsperger, M., et al., Conscious perception of errors and its relation to the anterior insula. Brain Struct Funct, 2010. 214(5-6): p. 629–43.10.1007/s00429-010-0261-1

53. Sutherland, M.T., et al., Resting state functional connectivity in addiction: Lessons learned and a road ahead. Neuroimage, 2012. 62(4): p. 2281–95.10.1016/j.neuroimage.2012.01.117

54. Menon, V. and L.Q. Uddin, Saliency, switching, attention and control: a network model of insula function. Brain Structure and Function, 2010. 214(5-6): p. 655–667

55. Klein, T.A., M. Ullsperger, and C. Danielmeier, Error awareness and the insula: links to neurological and psychiatric diseases. Front Hum Neurosci, 2013. 7: p. 14.10.3389/fnhum.2013.00014

56. Orr, C. and R. Hester, Error-related anterior cingulate cortex activity and the prediction of conscious error awareness. Front Hum Neurosci, 2012. 6: p. 177.10.3389/fnhum.2012.00177

57. Ramautar, J.R., et al., Probability effects in the stop-signal paradigm: the insula and the significance of failed inhibition. Brain Res, 2006. 1105(1): p. 143–54.10.1016/j.brainres.2006.02.091

58. O’Connell, R.G., et al., The neural correlates of deficient error awareness in attention-deficit hyperactivity disorder (ADHD). Neuropsychologia, 2009. 47(4): p. 1149–59.10.1016/j.neuropsychologia.2009.01.011

59. Goldstein, R.Z., et al., The neurocircuitry of impaired insight in drug addiction. Trends Cogn Sci, 2009. 13(9): p. 372–80.10.1016/j.tics.2009.06.004

60. Raichle, M.E., The brain’s default mode network. Annu Rev Neurosci, 2015. 38: p. 433-47.10.1146/annurev-neuro-071013-014030

61. Utevsky, A.V., D.V. Smith, and S.A. Huettel, Precuneus is a functional core of the default-mode network. J Neurosci, 2014. 34(3): p. 932–40.10.1523/JNEUROSCI.4227-13.2014

62. Fransson, P. and G. Marrelec, The precuneus/posterior cingulate cortex plays a pivotal role in the default mode network: Evidence from a partial correlation network analysis. Neuroimage, 2008. 42(3): p. 1178–84.10.1016/j.neuroimage.2008.05.059

63. Leech, R., et al., Fractionating the default mode network: distinct contributions of the ventral and dorsal posterior cingulate cortex to cognitive control. J Neurosci, 2011. 31(9): p. 3217–24.10.1523/JNEUROSCI.5626-10.2011

64. Anticevic, A., et al., The role of default network deactivation in cognition and disease. Trends Cogn Sci, 2012. 16(12): p. 584–92.10.1016/j.tics.2012.10.008

65. Li, C., et al., Greater activation of the “default” brain regions predicts stop signal errors. Neuroimage, 2007. 38(3): p. 640–648

66. Singh, K.D. and I.P. Fawcett, Transient and linearly graded deactivation of the human default-mode network by a visual detection task. Neuroimage, 2008. 41(1): p. 100–12.10.1016/j.neuroimage.2008.01.051

67. Allen, M., et al., The balanced mind: the variability of task-unrelated thoughts predicts error monitoring. Front Hum Neurosci, 2013. 7: p. 743.10.3389/fnhum.2013.00743

68. Gauffin, H., et al., Impaired language function in generalized epilepsy: inadequate suppression of the default mode network. Epilepsy Behav, 2013. 28(1): p. 26–35.10.1016/j.yebeh.2013.04.001

69. Whitfield-Gabrieli, S. and J.M. Ford, Default mode network activity and connectivity in psychopathology. Annu Rev Clin Psychol, 2012. 8: p. 49–76.10.1146/annurev-clinpsy-032511-143049

70. Oyegbile, T.O., et al., Default mode network deactivation in pediatric temporal lobe epilepsy: Relationship to a working memory task and executive function tests. Epilepsy Behav, 2019. 94: p. 124–130.10.1016/j.yebeh.2019.02.031

71. Bartova, L., et al., Reduced default mode network suppression during a working memory task in remitted major depression. J Psychiatr Res, 2015. 64: p. 9–18.10.1016/j.jpsychires.2015.02.025

72. Zhou, L., et al., Inefficient DMN Suppression in Schizophrenia Patients with Impaired Cognitive Function but not Patients with Preserved Cognitive Function. Sci Rep, 2016. 6: p. 21657.10.1038/srep21657

73. Zhang, R. and N.D. Volkow, Brain default-mode network dysfunction in addiction. Neuroimage, 2019. 200: p. 313–331.10.1016/j.neuroimage.2019.06.036

74. Bednarski, S.R., et al., Deficits in default mode network activity preceding error in cocaine dependent individuals. Drug Alcohol Depend, 2011. 119(3): p. e51–7.10.1016/j.drugalcdep.2011.05.026

75. Liddle, E.B., et al., Task-related default mode network modulation and inhibitory control in ADHD: effects of motivation and methylphenidate. J Child Psychol Psychiatry, 2011. 52(7): p. 761–71.10.1111/j.1469-7610.2010.02333.x

76. Peterson BS, et al., An FMRI study of the effects of psychostimulants on default-mode processing during Stroop task performance in youths with ADHD. American Journal of Psychiatry, 2009. 166(11): p. 1286–94

77. Seider, T.R., et al., Age exacerbates HIV-associated white matter abnormalities. J Neurovirol, 2016. 22(2): p. 201–12.10.1007/s13365-015-0386-3

78. Wendelken, L.A. and V. Valcour, Impact of HIV and aging on neuropsychological function. J Neurovirol, 2012. 18(4): p. 256–63.10.1007/s13365-012-0094-1

79. Valcour, V., et al., The Effects of Age and HIV on Neuropsychological Performance. J Int Neuropsychol Soc, 2011. 17(1): p. 190–5.10.1017/S1355617710001438

80. Saloner, R., et al., Effects of comorbidity burden and age on brain integrity in HIV. AIDS, 2019. 33(7): p. 1175–1185.10.1097/QAD.0000000000002192

81. Morgan, E.E., et al., Synergistic effects of HIV infection and older age on daily functioning. J Acquir Immune Defic Syndr, 2012. 61(3): p. 341–8.10.1097/QAI.0b013e31826bfc53

82. First, M.B., Structured clinical interview for the DSM (SCID). The encyclopedia of clinical psychology, 2014: p. 1–6

83. Broadbent, D., et al., The cognitive failures questionnaire (CFQ) and its correlates. British journal of clinical psychology, 1982. 21(1): p. 1–6

84. Larson, G. and C. Merritt, Can accidents be predicted? An empirical test of the Cognitive Failures Questionnaire. Applied Psychology, 1991. 40(1): p. 37–45

85. Albert, S.M., et al., Medication management skill in HIV: I. Evidence for adaptation of medication management strategies in people with cognitive impairment. II. Evidence for a pervasive lay model of medication efficacy. AIDS and Behavior, 2003. 7(3): p. 329–338

86. Albert, S.M., et al., An observed performance test of medication management ability in HIV: relation to neuropsychological status and medication adherence outcomes. AIDS, 1999. 3(2): p. 121–128

87. Patton, D.E., et al., Relationship of Medication Management Test-Revised (MMT-R) performance to neuropsychological functioning and antiretroviral adherence in adults with HIV. AIDS Behav, 2012. 16(8): p. 2286–96.10.1007/s10461-012-0237-7

88. Substance, A. and H.S.A. Mental, National Survey on Drug Use and Health: Summary of Methodological Studies. 2014: p. 1971–2014

89. Hester, R., et al., Neural mechanisms involved in error processing: a comparison of errors made with and without awareness. Neuroimage, 2005. 27(3): p. 602–8.10.1016/j.neuroimage.2005.04.035

90. Hester, R., et al., Neurochemical enhancement of conscious error awareness. J Neurosci, 2012. 32(8): p. 2619–27.10.1523/JNEUROSCI.4052-11.2012

91. Hester, R., C. Simoes-Franklin, and H. Garavan, Post-error behavior in active cocaine users: poor awareness of errors in the presence of intact performance adjustments. Neuropsychopharmacology, 2007. 32(9): p. 1974–84.10.1038/sj.npp.1301326

92. Ridderinkhof, K.R., et al., Neurocognitive mechanisms of cognitive control: the role of prefrontal cortex in action selection, response inhibition, performance monitoring, and reward-based learning. Brain Cogn, 2004. 56(2): p. 129–40.10.1016/j.bandc.2004.09.016

93. Esteban, O., et al., FMRIPrep: a robust preprocessing pipeline for functional MRI. bioRxiv, 2018: p. 306951.doi:10.1101/306951

94. Gorgolewski, K., et al., Nipype: a flexible, lightweight and extensible neuroimaging data processing framework in python. Front Neuroinform, 2011. 22(5): p. 13.10.3389/fninf.2011.00013

95. Abraham, A., et al., Machine learning for neuroimaging with scikit-learn. Frontiers in neuroinformatics, 2014. 8: p. 14

96. Tustison, N.J., et al., N4ITK: improved N3 bias correction. IEEE Trans Med Imaging., 2010. 29(6): p. 1310–1320.10.1109/TMI.2010.2046908

97. Fonov, V.S., et al., Unbiased nonlinear average age-appropriate brain templates from birth to adulthood. NeuroImage, 2009. 1(47): p. 102.10.1016/S1053-8119(09)70884-5

98. Jenkinson, M., et al., Improved optimization for the robust and accurate linear registration and motion correction of brain images. Neuroimage, 2002. 17(2): p. 825–841.10.1006/nimg.2002.1132

99. Cox, R.W., AFNI: software for analysis and visualization of functional magnetic resonance neuroimages. Comput Biomed Res, 1996. 29(3): p. 162–173.10.1006/cbmr.1996.0014

100. Huntenburg, J.M. Evaluating nonlinear coregistration of BOLD EPI and T1w images. 2014.

101. Wang, S., et al., Evaluation of Field Map and Nonlinear Registration Methods for Correction of Susceptibility Artifacts in Diffusion MRI. Front Neuroinform, 2017.10.3389/fninf.2017.00017

102. Treiber, J.M., et al., Characterization and Correction of Geometric Distortions in 814 Diffusion Weighted Image. PLoS One, 2016. 11(3).10.1371/journal.pone.0152472

103. Greve, D.N. and B. Fischl, Accurate and robust brain image alignment using boundary-based registration. xNeuroimage, 2009. 48(1): p. 63–72.10.1016/j.neuroimage.2009.06.060

104. Chen, G., P.A. Taylor, and R.W. Cox, Is the statistic value all we should care about in neuroimaging? NeuroImage, 2017. 147: p. 952–959.10.1016/j.neuroimage.2016.09.066

105. Cox, R.W., et al., FMRI Clustering in AFNI: False-Positive Rates Redux. Brain Connect, 2017. 7(3): p. 152–171.10.1089/brain.2016.0475

106. Hayes, A.F., Introduction to mediation moderation and conditional process analysis, in Methodology in the Social Sciences, D.A. Kenny, Editor. 2018, The Guilford Press: New York, London.

107. Shrout, P.E., Commentary: Mediation Analysis, Causal Process, and Cross-Sectional Data. Multivariate Behav Res, 2011. 46(5): p. 852–60.10.1080/00273171.2011.606718

108. Wechsler, D., Wechsler Test of Adult Reading: WTAR. Psychological Corporation, 2001

109. Fellows, R.P., D.A. Byrd, and S. Morgello, Effects of information processing speed on learning, memory, and executive functioning in people living with HIV/AIDS. J Clin Exp Neuropsychol, 2014. 36(8): p. 806–17.10.1080/13803395.2014.943696

110. Martin, E.M., et al., Stroop performance in drug users classified by HIV and hepatitis C virus serostatus. J Int Neuropsychol Soc, 2004. 10(2): p. 298–300.10.1017/S135561770410218X

111. Simmonds, D.J., J.J. Pekar, and S.H. Mostofsky, Meta-analysis of Go/No-go tasks demonstrating that fMRI activation associated with response inhibition is task-dependent. Neuropsychologia, 2008. 46(1): p. 224–32.10.1016/j.neuropsychologia.2007.07.015

112. Chevrier, A.D., M.D. Noseworthy, and R. Schachar, Dissociation of response inhibition and performance monitoring in the stop signal task using event-related fMRI. Hum Brain Mapp, 2007. 28(12): p. 1347–58.10.1002/hbm.20355

113. Wager, T.D., et al., Common and unique components of response inhibition revealed by fMRI. Neuroimage, 2005. 27(2): p. 323–40.10.1016/j.neuroimage.2005.01.054

114. Flannery, J., et al., Habenular and striatal activity during performance feedback is differentially linked with state-like and trait-like aspects of tobacco use disorder. Science Advances, 2019. 10(5): p. p.eaax2084

115. Ullsperger, M. and D.Y.v. Cramon, Error Monitoring Using External Feedback: Specific Roles of the Habenular Complex, the Reward System, and the Cingulate Motor Area Revealed by Functional Magnetic Resonance Imaging. J. Neurosci., 2003. 23(10): p. 4308 – 4314

116. Palminteri, S., et al., Critical roles for anterior insula and dorsal striatum in punishment-based avoidance learning. Neuron, 2012. 76(5): p. 998–1009.10.1016/j.neuron.2012.10.017

117. Craig, A.D., How do you feel--now? The anterior insula and human awareness. Nature reviews neuroscience, 2009. 10(1): p. 59–70

118. Cosentino, S., et al., The right insula contributes to memory awareness in cognitively diverse older adults. Neuropsychologia, 2015. 75: p. 163–9.10.1016/j.neuropsychologia.2015.05.032

119. Palaniyappan, L., et al., Appreciating symptoms and deficits in schizophrenia: right posterior insula and poor insight. Prog Neuropsychopharmacol Biol Psychiatry, 2011. 35(2): p. 523–7.10.1016/j.pnpbp.2010.12.008

120. Leech, R. and D.J. Sharp, The role of the posterior cingulate cortex in cognition and disease. Brain, 2014. 137(Pt 1): p. 12–32.10.1093/brain/awt162

121. Lou, H.C., J.P. Changeux, and A. Rosenstand, Towards a cognitive neuroscience of self-awareness. Neurosci Biobehav Rev, 2017. 83: p. 765–773.10.1016/j.neubiorev.2016.04.004

122. Weissman, D.H., et al., The neural bases of momentary lapses in attention. Nat Neurosci, 2006. 9(7): p. 971–8.10.1038/nn1727

123. Eichele, T., et al., Prediction of human errors by maladaptive changes in event-related brain networks. Proc Natl Acad Sci U S A, 2008. 105(16): p. 6173–8.10.1073/pnas.0708965105

124. Prado, J. and D.H. Weissman, Heightened interactions between a key default-mode region and a key task-positive region are linked to suboptimal current performance but to enhanced future performance. Neuroimage, 2011. 56(4): p. 2276–82.10.1016/j.neuroimage.2011.03.048

125. Hinds, O., et al., Roles of default-mode network and supplementary motor area in human vigilance performance: evidence from real-time fMRI. J Neurophysiol, 2013. 109(5): p. 1250–8.10.1152/jn.00533.2011

126. Li, C.S., et al., Greater activation of the “default” brain regions predicts stop signal errors. Neuroimage, 2007. 38(3): p. 640–8.10.1016/j.neuroimage.2007.07.021

127. Heilbronner, S.R. and M.L. Platt, Causal evidence of performance monitoring by neurons in posterior cingulate cortex during learning. Neuron, 2013. 80(6): p. 1384–91.10.1016/j.neuron.2013.09.028

128. Heaton, R.K., et al., The impact of HIV-associated neuropsychological impairment on everyday functioning. Journal of the International. Neuropsychological Society, 2004. 10(3): p. 317–331

129. Kamal, S., et al., The Presence of Human Immunodeficiency Virus-Associated Neurocognitive Disorders Is Associated With a Lower Adherence to Combined Antiretroviral Treatment. Open Forum Infect Dis, 2017. 4(2): p. ofx070.10.1093/ofid/ofx070

130. Meade, C.S., et al., Neurocognitive impairment and medication adherence in HIV patients with and without cocaine dependence. J Behav Med, 2011. 34(2): p. 128–38.10.1007/s10865-010-9293-5

131. Garrison, K.A., et al., Meditation leads to reduced default mode network activity beyond an active task. Cogn Affect Behav Neurosci, 2015. 15(3): p. 712–20.10.3758/s13415-015-0358-3

132. Salmi, J., L. Nyberg, and M. Laine, Working memory training mostly engages general-purpose large-scale networks for learning. Neurosci Biobehav Rev, 2018. 93: p. 108–122.10.1016/j.neubiorev.2018.03.019

133. Thames, A., et al., Depression, cognition, and self-appraisal of functional abilities in HIV: An examination of subjective appraisal versus objective performance. The Clinical Neuropsychologist, 2011. 25(2): p. 224–243

134. Garfield, S., et al., Suitability of measures of self-reported medication adherence for routine clinical use: a systematic review. BMC medical research methodology, 2011. 11(1): p. 1–9

135. Roberts, G.M. and H. Garavan, Evidence of increased activation underlying cognitive control in ecstasy and cannabis users. Neuroimage, 2010. 52(2): p. 429–35.10.1016/j.neuroimage.2010.04.192

136. Blest-Hopley, G., V. Giampietro, and S. Bhattacharyya, Regular cannabis use is associated with altered activation of central executive and default mode networks even after prolonged abstinence in adolescent users: Results from a complementary meta-analysis. Neurosci Biobehav Rev, 2019. 96: p. 45–55.10.1016/j.neubiorev.2018.10.026

137. Hinkin, C.H., et al., Medication adherence among HIV+ adults: effects of cognitive dysfunction and regimen complexity. Neurology, 2002. 59(12): p. 1944–1950

138. Spencer, S.J., M.P. Zanna, and G.T. Fong, Establishing a causal chain: why experiments are often more effective than mediational analyses in examining psychological processes. J Pers Soc Psychol, 2005. 89(6): p. 845–51.10.1037/0022-3514.89.6.845

139. O’Laughlin, K.D., M.J. Martin, and E. Ferrer, Cross-Sectional Analysis of Longitudinal Mediation Processes. Multivariate Behav Res, 2018. 53(3): p. 375–402.10.1080/00273171.2018.1454822

140. Maxwell, S.E., D.A. Cole, and M.A. Mitchell, Bias in cross-sectional analyses of longitudinal mediation: Partial and complete mediation under an autoregressive model. Multivariate Behavioral Research, 2011. 46(5): p. 816–841

