## Supplemental Information for "HIV infection is linked with reduced error-related default mode network suppression and poorer medication management abilities"

### SUPPLEMENTAL CONTENT

#### TABLES

- Table S1: Mental health status (pp. 2)
- Table S2: HIV status and immune system markers (pp. 3)
- Table S3: Plasma THC and metabolite levels (pp. 4)
- Table S4: Drug dependence (pp. 5)
- Table S5: Past month and lifetime drug use (pp. 6)
- Table S6: EAT trial type counts and percentages (pp. 7)
- Table S7: Cognitive control cluster coordinates (pp. 8)
- Table S8: Error awareness cluster coordinates (pp. 9)
- Table S9: Main effect of HIV cluster coordinates (pp. 10)

#### FIGURES AND LEGENDS

- Figure S1: Error awareness brain activity group effects. (pp. 11)
- Figure S2: Impact of cannabis use on brain activity. (pp. 12)

#### SUPPLEMENTAL REFERENCES

**SUPPLEMENTAL TABLES**

**Table S1. Participant mental health status by group.**

| | **All Participants** *n*=109 | **HIV+/CB+** *n*=32 | **HIV+/CB-** *n*=27 | **HIV-/CB+** *n*=28 | **HIV-/CB-** *n*=22 | **Group Effects (*p*'s)** HIV | CB |
|---|---|---|---|---|---|---|---|
| Major Depressive Episode (current) | 12 | 4 | 3 | 2 | 3 | 0.8 | 0.7 |
| Major Depressive Episode (past) | 25 | 11 | 3 | 4 | 7 | 0.8 | 0.6 |

NOTE. Mental health status is reported as count of participants meeting criteria for a current or past Major depressive episode as assessed via the Structured Clinical Interview for DSM-5 Research Version (SCID-5-RV) [1]. Participants were excluded from the sample if they presented severe mental illnesses with psychotic or paranoid symptoms. Group effects were assessed with two Chi-square tests, one comparing the HIV+ versus HIV- groups and the other comparing the CB+ versus CB- groups. No group differences were detected for any measure when conducting these same analyses after excluding participants who did not meet task engagement quality control threshold (n=2: HIV+/CB+; n=2: HIV+/CB-; n=2: HIV-/CB+).

**<u>Table S2.</u> Participant HIV status and immune system markers by group.**

| | **All HIV+ Participants** *n*=59 | **HIV+/CB+** *n*=32 | **HIV+/CB-** *n*=27 | **Group Effects (*p*'s)** |
|---|---|---|---|---|
| **Disease Characteristics** | | | | |
| Years since HIV diagnosis | 9.8 (9.2) | 8.7 (8.1) | 11.1 (10.3) | 0.3 |
| % with detectable viral load (>200 copies/mL) | 64.4 | 62.5 | 66.7 | 0.6 |
| % with AIDS diagnosis | 13.6 | 15.6 | 11.1 | 0.6 |
| **Viral Load (copies of HIV RNA/mL)** | | | | |
| HIV-1 (Linear) | 14,087.1 (54,905.1) | 18,654.9 (66,374.6) | 8,673.3 (37,628.1) | 0.2 |
| HIV-1 ($Log_{10}$) | 1.3 (1.7) | 1.4 (1.8) | 1.1 (1.5) | 0.2 |
| **Lymphocyte Subsets (cells/uL)** | | | | |
| T Helper Cells (CD3+CD4+) | 639.0 (304.4) | 647.6 (275.3) | 628.9 (340.9) | 0.8 |
| T Suppress./Cyt.(CD3+CB8+) | 1,064.8 (552.3) | 972.2 (426.0) | 1,174.5 (664.1) | 0.8 |

<u>NOTE</u>. Data are either expressed as mean (standard deviation) or as the percentage of participants. HIV-1 viral load assessed via the Abbott RealT*ime* HIV-1 assay. Group effects were assessed with independent samples *t*-tests or Chi-square tests comparing HIV+ cannabis users versus nonusers. No group differences were detected for any measure when conducting these same analyses after excluding participants who did not meet task engagement quality control threshold (n=2: HIV+/CB+ and n=2: HIV+/CB-).

**<u>Table S3.</u> Participant plasma THC and metabolite levels by group.**

| | **All CB+ Participants** *n*=60 | **HIV+/CB+** *n*=32 | **HIV-/CB+** *n* =28 | **Group Effects (*p*'s)** |
|---|---|---|---|---|
| 9-carboxy-THC (ng/mL) | 717.3 (1,087.1) | 853.1 (1,343.9) | 562.1 (679.7) | 0.3 |
| THC/creatinine ratio (ng/mg) | 596.8 (686.9) | 592.0 (712.0) | 602.6 (670.0) | 0.9 |

<u>NOTE</u>. Data are expressed as mean (standard deviation). Group effects were assessed with an independent samples *t*-test comparing cannabis using participants that were HIV+ versus HIV-. No group differences were detected for any measure when conducting these same analyses after excluding the participant who did not meet task engagement quality control threshold (n=2: HIV+/CB+ and n=2: HIV-/CB+).

**<u>Table S4.</u> Participant drug dependence status by group.**

| | **All Participants** *n*=109 | **HIV+/CB+** *n*=32 | **HIV+/CB-** *n*=27 | **HIV-/CB+** *n*=28 | **HIV-/CB-** *n*=22 | **Group Effects (*p*'s)** HIV | CB |
|---|---|---|---|---|---|---|---|
| % Cannabis (past month) | 10.1 | 18.8 | 0 | 17.9 | 0 | 0.9 | **0.002** |
| % Cannabis (lifetime) | 18.3 | 25.0 | 3.7 | 35.7 | 4.5 | 0.4 | **<0.001** |
| % Alcohol (lifetime) | 11.9 | 9.4 | 7.4 | 17.9 | 13.6 | 0.2 | 0.6 |
| % Cocaine (lifetime) | 11.0 | 9.4 | 7.4 | 14.3 | 13.6 | 0.4 | 0.8 |
| % Other stimulants (lifetime) | 3.7 | 9.4 | 3.7 | 0 | 0 | - | - |
| % Opiates (lifetime) | 1.8 | 3.1 | 0 | 3.6 | 0 | - | - |
| % Other sedatives (lifetime) | 1.8 | 6.3 | 0 | 0 | 0 | - | - |
| % Hallucinogens (lifetime) | 1.8 | 6.3 | 0 | 0 | 0 | - | - |
| % Other drug (lifetime) | 1.8 | 3.1 | 0 | 3.6 | 0 | - | - |

<u>NOTE</u>. Drug dependence data are expressed as the percentage of participants meeting dependency criteria either ever (lifetime) or in the past month (past month), as defined in the Structured Clinical Interview for DSM-5 Research Version (SCID-5-RV) [1]. As current (past month) drug dependence was exclusionary criteria (except cannabis dependence in the CB+ groups), no participants met dependency criteria in the past month for any of the reported drugs. Group effects were assessed with two Chi-square tests, one comparing the HIV+ vs. HIV- groups and one comparing the CB+ vs. CB- groups. These analyses were not performed when data did not meet criteria for Chi-square tests (cells with <5 observations). No significant differences were detected for any measure when conducting these same analyses after excluding participants who did not meet task engagement quality control threshold (n=2: HIV+/CB+; n=2: HIV+/CB-; n=2: HIV-/CB+).

**<u>Table S5.</u> Mean past month and lifetime participant drug use (times used).**

| | All Participants *n*=109 | HIV+/CB+ *n*=32 | HIV+/CB- *n*=27 | HIV-/CB+ *n*=28 | HIV-/CB- *n*=22 | Group Effects HIVxCB | HIV | CB |
|---|---|---|---|---|---|---|---|---|
| Cannabis (past month) | 13.0 (14.0) | 21.9 (10.7) | 0 | 25.6 (9.2) | 0 | *p*=0.2 | *p*=0.2 | ***p*<0.001*** |
| Cannabis (lifetime) | 2,404.7 (3,356.9) | 3,765.6 (3,488.9) | 114.0 (427.7) | 4,580.3 (3,733.1) | 467.5 (1,423.3) | *p*=0.7 | *p*=0.3 | ***p*<0.001*** |
| Alcohol (past month) | 2.3 (3.6) | 2.2 (2.7) | 2.1 (4.1) | 3.3 (4.6) | 1.6 (2.7) | *p*=0.2 | *p*=0.7 | *p*=0.2 |
| Alcohol (lifetime) | 1,204.7 (1,644.2) | 1,198.5 (1,634.7) | 1,184.8 (1,689.7) | 1,416.1 (1,537.1) | 969.2 (1,807.6) | *p*=0.5 | *p*=0.9 | *p*=0.5 |
| Cocaine (past month) | 0.0 (0.1) | 0.1 (0.2) | 0 | 0 | 0 | *p*=0.2 | *p*=0.2 | *p*=0.2 |
| Cocaine (lifetime) | 362.5 (1,186.0) | 351.0 (1,021.0) | 297.9 (1,026.4) | 312.0 (833.0) | 523.0 (1,860.8) | *p*=0.6 | *p*=0.7 | *p*=0.7 |
| Nicotine (past month) | 5.5 (11.3) | 8.2 (13.3) | 0.3 (1.5) | 7.9 (13.1) | 4.9 (10.8) | *p*=0.3 | *p*=0.3 | ***p*=0.012*** |
| Nicotine (lifetime) | 2,081.9 (3,731.2) | 2,765.7 (4,542.0) | 981.1 (2,540.5) | 2,366.3 (3,328.6) | 2,076.3 (4,065.8) | *p*=0.3 | *p*=0.6 | *p*=0.2 |
| Methamphetamine (past month) | 0.0 (0.2) | 0.1 (0.4) | 0 | 0 | 0 | *p*=0.4 | *p*=0.4 | *p*=0.4 |
| Methamphetamine (lifetime) | 25.0 (126.6) | 52.7 (208.8) | 27.6 (99.3) | 10.4 (54.4) | 0.05 (0.2) | *p*=0.8 | *p*=0.2 | *p*=0.5 |
| Prescription stimulants (past month) | 0.02 (0.2) | 0 | 0 | 0.1 (0.4) | 0 | *p*=0.3 | *p*=0.3 | *p*=0.3 |
| Prescription stimulants (lifetime) | 22.7 (207.2) | 72.8 (381.4) | 0 | 5.2 (22.7) | 0 | *p*=0.4 | *p*=0.4 | *p*=0.3 |
| Heroin (past month) | 0 | 0 | 0 | 0 | 0 | - | - | - |
| Heroin (lifetime) | 6.6 (69.0) | 0 | 0 | 25.8 (136.1) | 0.05 (0.2) | *p*=0.3 | *p*=0.3 | *p*=0.3 |
| Opiates (past month) | 0 | 0 | 0 | 0 (0.2) | 0 | - | - | - |
| Opiates (lifetime) | 17.8 (102.6) | 25.8 (127.2) | 0 | 39.3 (149.7) | 0.6 (2.5) | *p*=0.7 | *p*=0.7 | *p*=0.1 |
| Benzodiazepines (past month) | 0 | 0 | 0 | 0.04 (0.2) | 0 | *p*=0.3 | *p*=0.3 | *p*=0.3 |
| Benzodiazepines (lifetime) | 34.1 (252.7) | 112.9 (461.9) | 0 | 2.9 (7.3) | 1.1 (3.6) | *p*=0.3 | *p*=0.3 | *p*=0.2 |
| Barbiturates (past month) | 0 | 0 | 0 | 0 | 0 | - | - | - |
| Barbiturates (lifetime) | 6.5 (47.9) | 22.1 (87.4) | 0 | 0.04 (0.2) | 0 | *p*=0.2 | *p*=0.2 | *p*=0.2 |
| Ecstasy (past month) | 0 | 0 | 0 | 0 | 0 | - | - | - |
| Ecstasy (lifetime) | 27.6 (93.5) | 59.3 (147.9) | 0.5 (1.6) | 39.1 (84.6) | 0.1 (0.4) | *p*=0.6 | *p*=0.6 | ***p*=0.007*** |
| Inhalants (past month) | 0.04 (0.3) | 0.1 (0.5) | 0 | 0 | 0 | *p*=0.2 | *p*=0.2 | *p*=0.2 |
| Inhalants (lifetime) | 3.4 (17.0) | 57.2 (166.1) | 0.2 (1.0) | 2.5 (12.3) | 0 | *p*=0.1 | *p*=0.1 | *p*=0.1 |
| Ever injection drug use (count) | 2 | 1 | 0 | 1 | 0 | - | *p*=0.4 | *p*=0.4 |

<u>NOTE</u>. Data are expressed as mean (standard deviation). Drug use was self-reported as the number of "times" using each drug in the given timeframe (past month or lifetime) via items selected from the National Survey on Drug Use and Health [2]. Group effects were assessed with either an HIVxCB ANOVA or Chi square tests. * $p < 0.05$. No significant differences were detected for any measure when conducting these same analyses after excluding participants who did not meet task engagement quality control threshold (n=2: HIV+/CB+; n=2: HIV+/CB-; n=2: HIV-/CB+).

**<u>Table S6</u>. EAT trial type counts and percentages by sample (n=109, 103, or 86).**

| | **Trial Type Count** | ***n*=109** | | ***n*=103** | | ***n*=86** | |
|---|---|---|---|---|---|---|---|
| | | Mean ± SEM | Range | Mean ± SEM | Range | Mean ± SEM | Range |
| a) | Go-correct | 904.4 ± 15.6 | 275-1056 | 936.6 ± 9.3 | 588-1056 | 929.3 ± 10.9 | 588-1056 |
| | Go-error-omission | 63.5 ± 14.7 | 0-720 | 29.2 ± 5.4 | 0-313 | 34.4 ± 6.3 | 0-313 |
| b) | NoGo-correct | 115.7 ± 3.1 | 29-198 | 114.4 ± 3.1 | 29-198 | 114.6 ± 3.6 | 0-313 |
| | NoGo-error (commission) | 97.3 ± 3.3 | 18-187 | 98.5 ± 3.2 | 18-187 | 97.7 ± 3.7 | 18-187 |
| c) | NoGo-error-aware | 69.2 ± 3.1 | 0-134 | 71.9 ± 3.1 | 0-134 | 70.2 ± 3.2 | 3-122 |
| | NoGo-error-unaware | 21.1 ± 2.5 | 0-148 | 20.9 ± 2.6 | 0-148 | 21.2 ± 2.3 | 3-97 |
| | NoGo-error no response | 7.0 ± 1.4 | 0-123 | 5.8 ± 0.8 | 0-60 | 6.2 ± 0.9 | 0-60 |
| d) | NoGo-repeat-error | 40.7 ± 1.8 | 8-94 | 40.9 ± 1.8 | 8-94 | 41.6 ± 2.0 | 8-94 |
| | NoGo-repeat-error-aware | 24.9 ± 1.4 | 0-60 | 25.7 ± 1.4 | 0-60 | 25.2 ± 1.5 | 0-56 |
| | NoGo-repeat-error-unaware | 12.5 ± 1.5 | 0-80 | 12.4 ± 1.6 | 0-80 | 13.3 ± 1.7 | 0-80 |
| | NoGo-repeat-error no response | 3.3 ± 0.7 | 0-58 | 2.8 ± 0.4 | 0-34 | 3.1 ± 0.5 | 0-34 |
| e) | NoGo-Stroop-error | 56.7± 1.9 | 10-99 | 57.6 ± 1.9 | 10-99 | 56.0 ± 2.2 | 10-99 |
| | NoGo-Stroop-error-aware | 44.3 ± 2.0 | 0-82 | 46.2 ± 1.9 | 0-82 | 45.0 ± 2.0 | 2-82 |
| | NoGo-Stroop-error-unaware | 8.6 ± 1.2 | 0-82 | 8.4 ± 1.3 | 0-82 | 8.0 ± 1.0 | 0-51 |
| | NoGo-Stroop-error no response | 3.7 ± 0.7 | 0-65 | 3.0 ± 0.4 | 0-26 | 3.1 ± 0.4 | 0-26 |
| | **Trial Type Percent** | ***n*=109** | | ***n*=103** | | ***n*=86** | |
| | | Mean ± SEM | Range | Mean ± SEM | Range | Mean ± SEM | Range |
| a) | % Go-correct | 93.5 ± 1.5 | 30.1-100 | 96.9 ± 0.6 | 65.3-100 | 96.4 ± 6.2 | 65.3-100 |
| | % Go-error-omission | 6.5 ± 1.5 | 0-69.9 | 3.1 ± 0.6 | 0-34.7 | 3.7 ± 0.7 | 0-34.7 |
| b) | % NoGo-correct | 54.5 ± 1.5 | 13.4-91.7 | 53.9 ± 1.5 | 13.4-91.7 | 54.2 ± 1.7 | 13.4-91.7 |
| | % NoGo-error (commission) | 45.6 ± 1.5 | 8.3-86.6 | 46.1 ± 1.5 | 8.3-86.6 | 45.8 ± 1.7 | 8.3-86.6 |
| c) | % NoGo-error-aware | 72.0 ± 2.5 | 0-100 | 73.5 ± 2.4 | 0-100 | 72.5 ± 2.3 | 3.2-96.9 |
| | % NoGo-error-unaware | 21.7 ± 2.3 | 0-100 | 21.2 ± 2.3 | 0-100 | 21.8 ± 2.2 | 2.4-86.3 |
| | % NoGo-error no response | 6.3 ± 0.9 | 0-74.1 | 5.4 ± 0.7 | 0-33.5 | 5.8 ± 0.7 | 0-33.5 |
| d) | % NoGo-repeat-error | *41.4 ±1.6* | 9.8-74.6 | *40.9 ±1.0* | 19.8-74.6 | *42.1 ±1.0* | 21.6-74.6 |
| | % NoGo-repeat-error-aware | 64.6 ± 2.8 | 0-100 | 65.6 ± 2.8 | 0-100 | 63.1 ± 2.8 | 0-97.6 |
| | % NoGo-repeat-error-unaware | 28.8 ± 2.7 | 0-100 | 28.6 ± 2.8 | 0-100 | 30.6 ± 2.8 | 0-97.6 |
| | % NoGo-repeat-error no response | 6.6 ± 1.0 | 0-77.3 | 5.8 ± 0.7 | 0-42.5 | 6.3 ± 0.3 | 0-42.5 |
| e) | % NoGo-Stroop-error | *58.6 ±1.7* | 25.4-80.2 | *59.1 ±1.0* | 25.4-80.2 | *57.9 ±1.0* | 25.4-78.4 |
| | % NoGo-Stroop-error-aware | 77.6 ± 2.4 | 0-100 | 79.3 ± 2.3 | 0-100 | 79.5 ± 2.1 | 12.5-100 |
| | % NoGo-Stroop-error-unaware | 16.2 ± 2.1 | 0-100 | 15.5 ± 2.1 | 0-100 | 15.0 ± 1.8 | 0-75 |
| | % NoGo-Stroop-error no response | 6.2 ± 0.9 | 0-71.4 | 5.2 ± 0.7 | 0-39.3 | 5.5 ± 0.7 | 0-39.3 |

<u>NOTE</u>. The *n*=103 sample was used for error-related brain and behavior assessments and excluded 6 participants from the total sample (*n*=109) that did not meet task performance criteria (more than 50% Go-errors). The *n*=86 sample was used for error-awareness-related brain and behavior assessments and excluded an additional 17 participants that did not commit at least 2 of each type of NoGo-error (aware and unaware). **<u>Trial Type Count</u>**: Rows in section a) do not add up to 1080 total Go trails because for every incorrect NoGo trial, an Awareness-trial, in which participants are instructed to press button-2 to indicate error awareness, takes the place of the Go trial immediately following the error. Further, four subjects only completed 4 out of the 6 task runs (720 total Go trails and 144 total NoGo trials) and one subject only completed 5 out of the 6 task runs (900 total Go trials and 180 total NoGo trials). This is also why rows in section b) do not add up to 216 total NoGo trials. **<u>Trial Type Percent</u>:** This section reports the **<u>a)</u>** percentage of correct and error Go trails out of total Go trials (adds up to 100%) and **<u>b)</u>** the percentage of correct and error NoGo trials out of the total NoGo trials (adds up to 100%). **<u>c)</u>** The percentage of aware, unaware and no response NoGo-errors out of all NoGo-errors (adds up to 100%). **<u>d-e)</u>** NoGo-repeat-errors and NoGo-Stroop-errors out of all NoGo errors (values italicized) such that these italicized values add up to 100%.

**Table S7. Cognitive control success (inhibition) and failure (error) cluster coordinates.** Regions showing greater activation during successful (NoGo-correct) and failed (NoGo-error) NoGo trials. Whole-brain, one-sample *t*-test (two-tailed, 3dTtest++), thresholded at $p_{voxel\text{-}wise}$=1.0e$^{-10}$, cluster extent: 20 voxels (arbitrarily chosen). ***n*=103**: 6 participants from the total sample (*n*=109) were excluded because they did not meet task performance criteria (more than 50% Go-errors).

| | Region | Hemisphere | Center Coordinates (MNI, LPI) | | | Cluster Size (# of Voxels) |
|---|---|---|---|---|---|---|
| | | | X | Y | Z | |
| **NoGo-Correct (C > E)** | | | | | | |
| 1 | **Middle occipital** gyrus | R | 32 | -79 | 15 | 625 |
| 2 | **Middle occipital** gyrus | L | -29 | -85 | 13 | 564 |
| 3 | **Cerebral white matter** | L | -27 | -24 | 33 | 236 |
| 4 | **Ventral striatum** (putamen, nucleus accumbens) | R | 22 | 11 | -4 | 116 |
| 5 | **Paracentral** gyrus (primary motor cortex, primary sensory cortex) | R | 10 | -30 | 65 | 104 |
| 6 | **Superior temporal** gyrus | R | 61 | -15 | 2 | 64 |
| 7 | **Ventral striatum** (putamen, nucleus accumbens) | L | -22 | 10 | -6 | 49 |
| 8 | **Paracentral** gyrus (primary motor cortex, primary sensory cortex) | L | -10 | -31 | 64 | 45 |
| 9 | **Hippocampus** (parahippocampus, amygdala) | R | 25 | -9 | -18 | 43 |
| 10 | **Cerebral white matter** | R | 25 | 29 | 18 | 40 |
| 11 | **Hippocampus** (parahippocampus, amygdala) | L | -27 | -11 | -17 | 33 |
| 12 | **Cerebral white matter** | R | 26 | 1 | 35 | 20 |
| **NoGo-Error (E > C)** | | | | | | |
| 1 | **Postcentral** gyrus/inferior parietal gyrus | L | -46 | -27 | 54 | 814 |
| 2 | **SMA/Pre-SMA/**superior, middle & medial frontal gyrus | B | -1 | 18 | 48 | 223 |
| 3 | **Cerebellum** (culmen & dentate) | R | 19 | -55 | -21 | 144 |
| 4 | **Anterior Insula**/inferior frontal gyrus (IFG) | L | -46 | 18 | -2 | 106 |
| 5 | **Thalamus** | L | -10 | -23 | 8 | 75 |
| 6 | **Precentral** gyrus | L | -56 | 8 | 31 | 66 |
| 7 | **Cerebellum** | R | 17 | -66 | -54 | 57 |
| 8 | **Anterior Insula**/inferior frontal gyrus (IFG) | R | 53 | 18 | 0 | 41 |
| 9 | **Postcentral** gyrus | R | 61 | -16 | 35 | 34 |
| 10 | **Middle insula**/frontal opercular area | L | -45 | -4 | 12 | 21 |

NOTE. Voxel size: 3.44 x 3.44 x 3.40 mm$^3$. X: Left (-), Right (+); Y: Posterior (-), Anterior (+); Z: Inferior (-), Superior (+). Region labels come from the AFNI Talairach daemon atlas. See main text Figure 2A for graphical representation.

**<u>Table S8.</u> Error awareness cluster coordinates.** Regions showing greater activation during aware and unaware errors on NoGo trials. Whole-brain, one-sample *t*-test on NoGo-error: Aware minus NoGo-error: Unaware [A-U] contrast values ($p_{FWE\text{-}corrected}$<0.05; $p_{voxel\text{-}wise}$<0.0001, cluster extent: 7 voxels, 3dClustSim with spatial autocorrelation correction [3]). ***n*=86**: 23 participants from the total sample that either did not meet task performance criteria (more than 50% Go-errors) or did not commit at least 2 of each type of NoGo-error (aware and unaware) were excluded.

| | Region | Hemisphere | Center Coordinates (MNI, LPI) | | | Cluster Size (# of Voxels) |
|---|---|---|---|---|---|---|
| | | | X | Y | Z | |
| **Aware NoGo-errors (A > U)** | | | | | | |
| 1 | **Postcentral, Precentral & inferior parietal** gyrus | B | -38 | 25 | 72 | 2330 |
| 2 | **Cerebellum** (vermis) | B | 0 | 46 | 0 | 1440 |
| 3 | **Thalamus, putamen, posterior insula** | R | 0 | 39 | 4 | 586 |
| 4 | **Postcentral** gyrus | R | 45 | 39 | 65 | 487 |
| 5 | **Anterior insula**/inferior frontal gyrus | R | 59 | -16 | -3 | 179 |
| 6 | **Middle cingulate** cortex | B | 0 | 29 | 27 | 92 |
| 7 | **Ventral striatum** (putamen, nucleus accumbens) | R | 28 | -2 | -10 | 71 |
| 8 | **Superior parietal** gyrus (precuneus) | R | 7 | 77 | 55 | 45 |
| 9 | **Superior frontal** gyrus | R | 35 | -54 | 31 | 39 |
| 10 | **Superior frontal** gyrus | L | -34 | -54 | 27 | 34 |
| 11 | **Middle posterior temporal** gyrus | R | 66 | 53 | 4 | 7 |
| **Unaware NoGo-errors (U > A)** | | | | | | |
| 1 | **Middle frontal** gyrus | R | 28 | -30 | 58 | 40 |
| 2 | **Inferior frontal** gyrus (orbitalis) | R | 42 | -40 | -17 | 18 |
| 3 | **Superior frontal** gyrus | L | -24 | -37 | 48 | 18 |
| 4 | **Inferior frontal** gyrus (triangularis) | R | 52 | -26 | 14 | 12 |
| 5 | **Posterior cingulate** cortex | R | 11 | 56 | 14 | 11 |
| 6 | **Superior parietal** gyrus (precuneus) | L | -48 | -77 | 41 | 10 |
| 7 | **Posterior cingulate** cortex | L | -10 | -56 | 10 | 8 |

<u>NOTE.</u> Voxel size: 3.44 x 3.44 x 3.40 $mm^3$. X: Left (-), Right (+); Y: Posterior (-), Anterior (+); Z: Inferior (-), Superior (+). Region labels come from the AFNI Talairach daemon atlas. See main text Figure 3A for graphical representation.

**Table S9.** **Main effect of HIV serostatus cluster coordinates.** Regions showing significant HIV main effects in a whole-brain, HIV x CB ANOVA (3dMVM) assessing cognitive-control/failure [C-E] activation maps ($p_{FWE\text{-}corrected}$<0.05; $p_{voxel\text{-}wise}$<0.001, cluster extent: 35 voxels). ***n*=103**: 6 participants from the total sample that did not meet task performance criteria (more than 50% Go-errors) were excluded.

| | Region | Hemisphere | Center Coordinates (MNI, LPI) X | Y | Z | Cluster Size (# of Voxels) |
|---|---|---|---|---|---|---|
| **HIV main effect** | | | | | | |
| 1 | **Medial prefrontal** cortex (mPFC) | B | 2 | 65 | -2 | 39 |
| 2 | **Posterior cingulate** cortex (PCC) | B | 2 | -51 | 29 | 36 |

NOTE. Voxel size: 3.44 x 3.44 x 3.40 $mm^3$. X: Left (-), Right (+); Y: Posterior (-), Anterior (+); Z: Inferior (-), Superior (+). Region labels come from the AFNI Talairach daemon atlas. See main text Figure 4A for graphical representation.

### SUPPLEMENTAL FIGURES AND LEGENDS

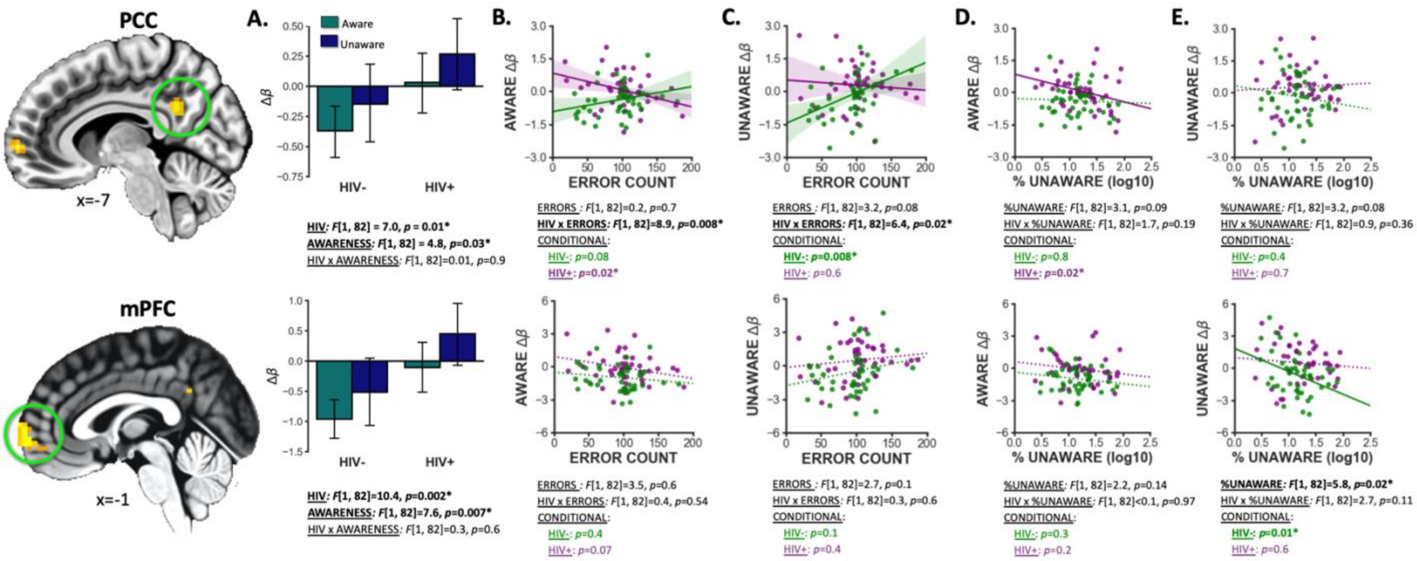


**Figure S1. Error awareness brain activity in HIV-positive and HIV-negative groups.** At the whole-brain level, no significant group effects on aware vs. unaware error [A-U] contrast images were detected. However, follow-up exploratory analyses on average [A] and [U] $\beta$ coefficients extracted from PCC and mPFC ROI masks were conducted. **(A)** 3-way 2(HIV: positive vs. negative) x 2(CB: user vs, nonuser) x 2(AWARENESS: aware vs. unaware) ANOVAs were conducted separately for each ROI (*n*=86) Similar to the inhibition analyses reported in the main text (**main text Fig. 4A**), no significant HIV x CB interaction or CB main effects were observed; however, both the PCC ($F[1, 82] = 4.8$, $p$=0.03) and mPFC ($F[1, 82] = 7.6$, $p$=0.007) displayed a significant main effect of AWARENESS such that both regions displayed increased deactivation during aware errors relative to unaware errors. Further, consistent with the HIV main effect on brain deactivation during all errors (**main text Fig. 4A**), both regions also displayed a significant main effect of HIV such that, HIV- controls had increased deactivation in the PCC ($F[1, 82] = 7.0$, $p = 0.01$) and mPFC ($F[1, 82] = 10.4$, $p = 0.002$) compared to the HIV+ individuals. While these results indicate significantly less PCC and mPFC deactivation among the HIV+ group during both aware and unaware errors, the nonsignificant HIV x AWARENESS interactions ($p$'s>0.6) denote that both HIV- and HIV+ groups display a similar difference between aware and unaware error brain activity. (**B: *n*=86**) When assessing relations to task behavior we observed significant HIV by ERROR-COUNT interactions on PCC activity during aware errors ($F[1, 82]$=8.9, $P_{Bonferronni\text{-}corrected} = 0.008$) similar to interaction effects on PCC activity during all errors that was reported in the main text (**main text Fig. 4C)**. Follow-up conditional effects within each group indicated that, among HIV+ participants, increased PCC deactivation during aware errors was significantly associated with worsened cognitive control task performance (more NoGo-errors; $p$=0.02). (**C: n=86**) We also observed significant HIV by ERROR-COUNT interactions on PCC activity during and unaware errors ($F[1, 82]$=6.4, $P_{Bonferronni\text{-}corrected} = 0.02$). Follow-up conditional effects within each group indicated that, among HIV- controls, increased PCC deactivation during unaware errors was associated with better cognitive control task performance (fewer NoGo-errors) whereas, among HIV+ participants, increased PCC activity during aware errors was associated with worsened cognitive control task performance. (**D: *n*=86**) We observed null effects of behavioral error awareness performance (%UNAWARE) on brain activity during aware errors ($p$'s>0.1). (**E: *n*=86**) We observed a significant main effect of awareness performance on mPFC brain activity during unaware errors such that decreased mPFC activity during unaware errors was linked to a higher percent of unaware errors ($F[1, 82]$=5.8, $P_{Bonferronni\text{-}corrected} = 0.02$). Follow-up conditional effects within each group indicated that this main effect of error awareness performance on mPFC unaware activity was primarily displayed among HIV- controls ($p$=0.01).

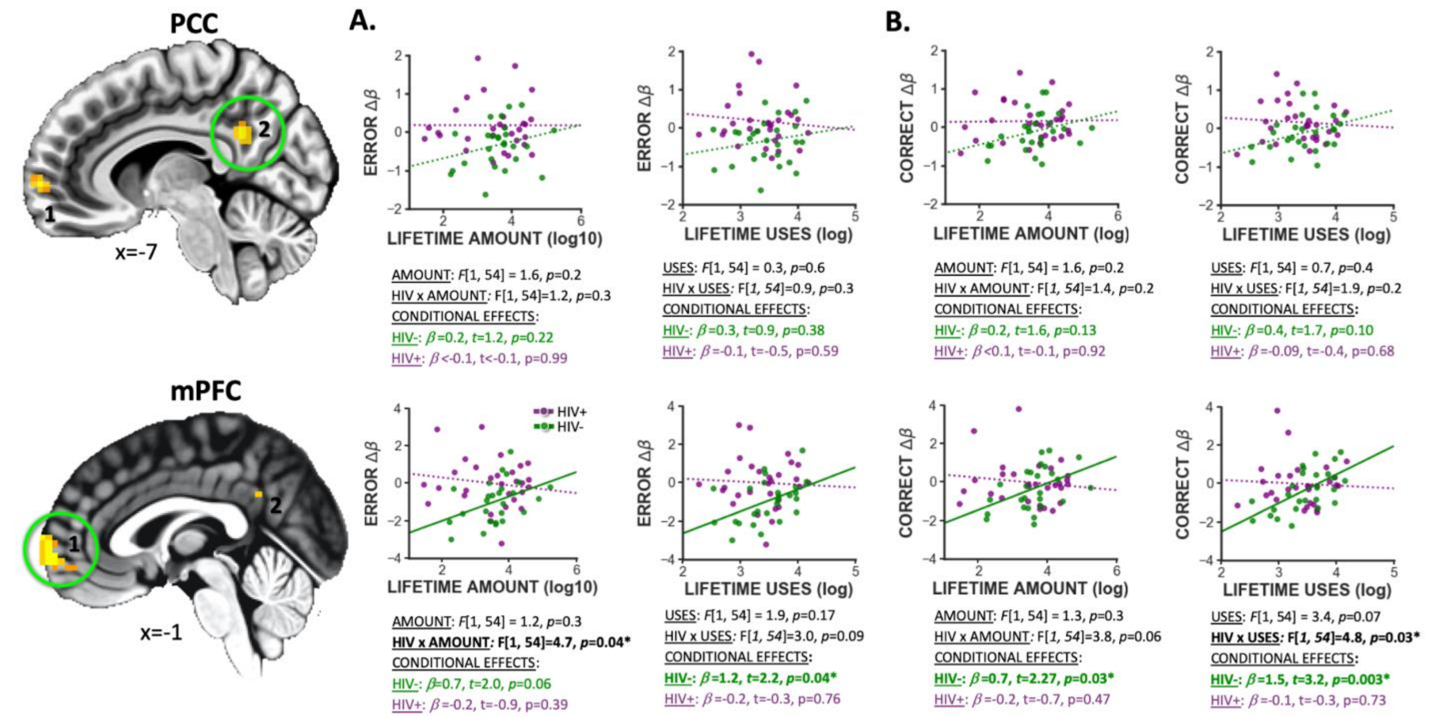


**Figure S2. Impact of chronic CB use on brain activity.** To assess the impact of self-reported amount of CB used over the lifetime (LIFETIME-AMOUNT) and the impact of total times using CB (LIFETIME-USES) on HIV-associated functional brain alterations, we conducted exploratory HIV x LIFETIME-AMOUNT and HIV x LIFETIME-USES ANCOVAs on task-related $\beta$s (NoGo-error and NoGo-correct) averaged across voxels in ROIs displaying a significant main effect of HIV, among CB using participants ($n$=55). Past month amount of nicotine used and past month times using nicotine variables were included as covariates in these analyses. As the LIFETIME-AMOUNT and LIFETIME-USE variables were skewed to the right, both were $\log_{10}$ transformed before conducting statistical tests. **(A)** During error-NoGo trials: among HIV- users, but not PLWH users, more times using CB over the lifetime ($\beta$=1.2, $t[24]$=2.2, $p$=0.04) was significantly linked to reduced deactivation in the mPFC. A similar, albeit nonsignificant pattern was observed for HIV- controls' amount of cannabis used over the lifetime ($\beta$=0.7, $t[24]$=2.0, $p$=0.06). (**B**) During correct-NoGo trials: among HIV- CB users, but not PLWH users, both times using CB over the lifetime ($\beta$=1.5, $t[24]$=3.2, $p$=0.003) and amount of CB used over the lifetime ($\beta$=0.7, $t[24]$=2.3, $p$=0.03) were significantly associated with reduced deactivation in the mPFC. No significant effects of LIFETIME-USES or LIFETIME-AMOUNT on PCC activity were observed.

### SUPPLEMENTAL REFERENCES

1. First, M., et al., Structured clinical interview for DSM-5—Research version (SCID-5 for DSM-5, research version; SCID-5-RV). Arlington, VA. *American Psychiatric Association*, 2015
2. Substance, A. and H.S.A. Mental, National Survey on Drug Use and Health: Summary of Methodological Studies. 2014: p. 1971–2014
3. Cox, R.W., et al., FMRI Clustering in AFNI: False-Positive Rates Redux. *Brain Connect*, 2017. **7**(3): p. 152-171.10.1089/brain.2016.0475
